# Validation of a Multivariate Prediction Model of the Clinical Progression of Alzheimer’s Disease in a Community-Dwelling Multiethnic Cohort

**DOI:** 10.1101/2022.06.28.22277006

**Authors:** Eric Stallard, Anton Kociolek, Zhezhen Jin, Hyunnam Ryu, Seonjoo Lee, Stephanie Cosentino, Carolyn Zhu, Yian Gu, Kayri Fernandez, Michelle Hernandez, Bruce Kinosian, Yaakov Stern

## Abstract

**Background:** The major aims of the three Predictors Studies have been to further our understanding of Alzheimer’s disease (AD) progression sufficiently to predict the length of time from disease onset to major disease outcomes in individual patients with AD.

**Objectives:** To validate a longitudinal Grade of Membership (L-GoM) prediction algorithm developed using clinic-based, mainly white patients from the Predictors 2 Study in a statistically representative community-based sample of Hispanic (*N*=211) and non-Hispanic (*N*=62) older adults from the Predictors 3 Study and extend the algorithm to mild cognitive impairment (MCI).

**Methods:** The L-GoM model was applied to data collected at the initial Predictors 3 visit for 150 subjects with AD and 123 with MCI. Participants were followed annually for up to seven years. Observed rates of survival and need for full-time care (FTC) were compared to those predicted by the algorithm.

**Results:** Initial MCI/AD severity in Predictors 3 was substantially higher than among clinic-based AD patients enrolled at the specialized Alzheimer’s centers in Predictors 2. The observed survival and need for FTC followed the L-GoM model trajectories in individuals with MCI or AD, except for *N*=32 subjects initially diagnosed with AD who reverted to a non-AD diagnosis on follow-up.

**Conclusions:** These findings indicate that the L-GoM model is applicable to community-dwelling, multiethnic older adults with AD. They extend the use of the model to the prediction of outcomes for MCI. They also justify release of our L-GoM calculator at this time.

## INTRODUCTION

The course of Alzheimer’s disease varies markedly across patients. We recently introduced a model of the progression of Alzheimer’s disease (AD) that uses a longitudinal Grade of Membership (L-GoM) approach to model disease progression [1]. Developed using data from the Predictors 2 Study, the L-GoM model incorporates measures of 11 key domains (see Table 1) at each patient’s initial visit and every six months for up to 10 years. The L-GoM model can use data from an individual at any point in the disease process to predict the likely future trajectory for that individual. We subsequently validated the model on a separate data set from the Predictors 1 Study [2], showing that the model made accurate predictions of time to important disease outcomes.

**Table 1.** Domains of measurement, instruments, and descriptions of covariates used in the longitudinal Grade of Membership model.

| Domain | Instrument | Variable Names |
| --- | --- | --- |
| **Fixed Covariates (N=6)** |  |  |
| Baseline characteristics | Initial assessment | ApoE genotype, sex, age at intake, ethnicity/race, occupation, *years since diagnosis* |
| **Time-Varying Variables (N=73)** |  |  |
| Acute medical treatments/conditions | Patient follow-up questionnaire | *Admission to hospital, treatment, had seizure?* |
| Alcohol use | Alcohol questionnaire | *Beer/week, wine/week, hard liquor/week* |
| Behavior | CUSPAD [33] | Wandering away, verbal outbursts, physical threats, difficulty sleeping |
| Cognition | MMSE [34] | Orientation, registration, “world” backward, recall, language, drawing; *MMSE completion indicator* |
| Dementia with Lewy body symptoms | DLB questionnaire [35] | Fluctuating cognition, visual hallucinations; *DLB questionnaire completion indicator* |
| Dependence | Dependence scale [11] | Dependence scale (13 items), equivalent institutional care, type of residence/facility, *length of stay in LTC facility* |
| Depression/agitation | CUSPAD [33] | Agitation, sadness/depression, depression frequency, appetite problems |
| Eyesight/hearing | Medical questionnaire | Adequate sight? Adequate hearing? |
| Function, Personality | BDRS [36] | IADL (8 items), BADL (3 items), *personality (11 items)* |
| Motor signs/symptoms | UPDRS [37] | Extrapyramidal signs (summary score), tremor, bradykinesia, gait, myoclonus, rigidity |
| Psychiatric/psychotic symptoms | CUSPAD [33] | Delusions, hallucinations, illusions |

**Notes:** 21 variables with names listed in italics were not used in the present validation study; the remaining 58 variables were used to estimate the initial L-GoM scores in Predictors 3 using procedures for randomly missing data described in [2] to handle the approximately 5.7% of missing data points.
**Abbreviations:** ApoE Apolipoprotein E, BADL Basic activities of daily living, BDRS Blessed Dementia Rating Scale, CUSPAD Columbia University Scale for Psychopathology in Alzheimer’s Disease, DLB Dementia with Lewy bodies, IADL Instrumental activities of daily living, MMSE Mini Mental State Examination, UPDRS Unified Parkinson’s Disease Rating Scale.

The Predictors 1 and 2 data sets used to derive and validate the L-GoM model both consisted of clinic-based, non-Hispanic white patients with high socioeconomic status (SES), all initially with mild AD. In addition, all were patients at specialized Alzheimer’s centers. It is important to determine whether or not the model operates equally well among community-dwelling older adults of different ethnic, linguistic, and SES backgrounds.

The Predictors 3 cohort was developed to examine this issue [3]. This cohort consists of community-dwelling, multiethnic older adults in the Washington Heights and Inwood areas of Manhattan, New York who were assigned research diagnoses of prevalent or incident AD, as well as individuals whose cognitive or functional status suggested they were at risk for converting to AD in the near future—individuals who were assigned research diagnoses of mild cognitive impairment (MCI). In order to validate the L-GoM model in this cohort, we applied the model to information acquired at the first dementia or MCI visit—i.e., the initial visit of subjects diagnosed with prevalent AD dementia, the visit at which incident cases were diagnosed with AD dementia, or the visit at which MCI was first detected. We used the L-GoM model [1] to predict individual-specific survival probabilities over the first 7.5 years after the initial visit, and individual-specific prevalence probabilities of the need for full-time care (FTC) at the initial visit and each year thereafter. We aggregated the individual subjects into various diagnostic and demographic groups and subgroups in order to validate the L-GoM predicted probabilities against the corresponding group-specific observed proportions.

Validation of the L-GoM model in the Predictors 3 cohort can extend its utility in important ways by establishing its applicability to community-dwelling, multiethnic older adults with AD in addition to patients recruited from a clinical setting. Predictors 3 also allows us to explore the broader application of the L-GoM model to individuals with MCI. These applications can help establish the model as an important predictive tool for individuals with AD or MCI, clinicians, clinical trialists, and other researchers.

## METHODS

### The Predictors 3 Cohort

Community-based subjects for the Predictors 3 cohort were recruited from the Washington Heights Inwood Columbia Aging Project (WHICAP). WHICAP participants were first identified from a random sample of elderly Medicare recipients residing in the designated areas of Washington Heights, Hamilton Heights, and Inwood in North Manhattan, NY. Potential participants were excluded at the time of recruitment if they did not speak English or Spanish. WHICAP participants were evaluated approximately every 18 months. WHICAP follows a combination of remaining participants from cohorts originally recruited in 1992 (N = 2,332), 1999–2001 (N = 2,776), and 2009–2015 (N = 2,088). Recruitment of the Predictors 3 cohort began in 2011 and continued to 2019. The primary goal was to recruit subjects from WHICAP’s incident and prevalent AD dementia cases. The diagnosis of AD dementia was based on the 2011 criteria [4]; the Clinical Dementia Rating Scale (CDR) [5] was used to rate dementia severity. We also enrolled WHICAP participants with MCI. These included participants who received the diagnosis of MCI using an implementation of MCI criteria [6, 7] consistent with the 2011 criteria [8]. These also included others who received a CDR score of 0.5 based on the WHICAP algorithm for evaluating neuropsychological test scores [9] and activities of daily living. All participants’ data were reviewed in a consensus conference at each follow-up assessment, at which time the conferees ascertained if/when any at-risk individuals with MCI converted to AD dementia, and whether those diagnosed with AD retained that diagnosis. Except as noted in the next paragraph, the present study included all AD cases, whether or not they reverted to a non-dementia diagnosis, and similarly all MCI cases, whether or not they converted to AD dementia at a subsequent follow-up visit.

Once included in the Predictors 3 Study, subjects were followed annually using many of the same instruments administered in Predictors 2 [3], including all instruments in Table 1. Of the 292 participants, 19 were lost to follow-up after the first visit; they were excluded from the present study because they did not contribute to our longitudinal analysis. Baseline characteristics of the 273 subjects used for model validation are displayed in Table 2 along with comparable statistics for the 229 subjects used for model development in Predictors 2.

**Table 2.** Characteristics of Predictors 2 and 3 Cohorts at Baseline Visit.

|  | Predictors 2 |  | Predictors 3 |  |  |  |  |  |  |  |  |  |
| --- | --- | --- | --- | --- | --- | --- | --- | --- | --- | --- | --- | --- |
|  | Full Sample |  | Full Sample |  | MCI |  |  |  | AD Dementia |  |  |  |
|  |  |  |  |  | Nonconverters |  | Converters |  | Reverters |  | Nonreverters |  |
|  | N | % (Excl. Unkn.) | N | % (Excl. Unkn.) | N | % (Excl. Unkn.) | N | % (Excl. Unkn.) | N | % (Excl. Unkn.) | N | % (Excl. Unkn.) |
| Total | 229 | 100% | 273 | 100% | 83 | 100% | 40 | 100% | 32 | 100% | 118 | 100% |
| Sex |  |  |  |  |  |  |  |  |  |  |  |  |
| Male | 91 | 39.7% | 60 | 22.0% | 23 | 27.7% | 11 | 27.5% | 6 | 18.8% | 20 | 16.9% |
| Female | 138 | 60.3% | 213 | 78.0% | 60 | 72.3% | 29 | 72.5% | 26 | 81.3% | 98 | 83.1% |
| Age |  |  |  |  |  |  |  |  |  |  |  |  |
| 49-64 | 20 | 8.7% | - | 0.0% | - | 0.0% | - | 0.0% | - | 0.0% | - | 0.0% |
| 65-69 | 14 | 6.1% | - | 0.0% | - | 0.0% | - | 0.0% | - | 0.0% | - | 0.0% |
| 70-74 | 48 | 21.0% | 22 | 8.1% | 10 | 12.0% | 2 | 5.0% | 4 | 12.5% | 6 | 5.1% |
| 75-79 | 69 | 30.1% | 29 | 10.6% | 9 | 10.8% | 2 | 5.0% | 6 | 18.8% | 12 | 10.2% |
| 80-84 | 46 | 20.1% | 72 | 26.4% | 19 | 22.9% | 12 | 30.0% | 6 | 18.8% | 35 | 29.7% |
| 85+ | 32 | 14.0% | 150 | 54.9% | 45 | 54.2% | 24 | 60.0% | 16 | 50.0% | 65 | 55.1% |
| Ethnicity/Race |  |  |  |  |  |  |  |  |  |  |  |  |
| Hispanic | 8 | 3.6% | 211 | 77.3% | 55 | 66.3% | 29 | 72.5% | 28 | 87.5% | 99 | 83.9% |
| Non-Hispanic White | 202 | 90.2% | 25 | 9.2% | 13 | 15.7% | 6 | 15.0% | - | 0.0% | 6 | 5.1% |
| Non-Hispanic Nonwhite | 14 | 6.3% | 37 | 13.6% | 15 | 18.1% | 5 | 12.5% | 4 | 12.5% | 13 | 11.0% |
| Unknown | 5 | 2.2% | - | 0.0% | - | 0.0% | - | 0.0% | - | 0.0% | - | 0.0% |
| Education (Years) |  |  |  |  |  |  |  |  |  |  |  |  |
| 0-8 | 8 | 3.5% | 191 | 70.0% | 59 | 71.1% | 23 | 57.5% | 24 | 75.0% | 85 | 72.0% |
| 9-12 | 85 | 37.1% | 47 | 17.2% | 11 | 13.3% | 8 | 20.0% | 5 | 15.6% | 23 | 19.5% |
| 13+ | 136 | 59.4% | 35 | 12.8% | 13 | 15.7% | 9 | 22.5% | 3 | 9.4% | 10 | 8.5% |
| APOE Genotype |  |  |  |  |  |  |  |  |  |  |  |  |
| e4/e4 | 20 | 11.8% | 5 | 1.8% | 1 | 1.2% | 2 | 5.0% | 1 | 3.1% | 1 | 0.8% |
| e3/e4 | 71 | 42.0% | 74 | 27.1% | 15 | 18.1% | 13 | 32.5% | 4 | 12.5% | 42 | 35.6% |
| e2/e4 | 3 | 1.8% | 6 | 2.2% | 1 | 1.2% | 1 | 2.5% | - | 0.0% | 4 | 3.4% |
| e3/e3 | 62 | 36.7% | 156 | 57.1% | 58 | 69.9% | 20 | 50.0% | 21 | 65.6% | 57 | 48.3% |
| e2/e3 | 13 | 7.7% | 32 | 11.7% | 8 | 9.6% | 4 | 10.0% | 6 | 18.8% | 14 | 11.9% |
| Unknown | 60 | 26.2% | - | 0.0% | - | 0.0% | - | 0.0% | - | 0.0% | - | 0.0% |
| Any APOE e4? |  |  |  |  |  |  |  |  |  |  |  |  |
| No | 75 | 44.4% | 188 | 68.9% | 66 | 79.5% | 24 | 60.0% | 27 | 84.4% | 71 | 60.2% |
| Yes | 94 | 55.6% | 85 | 31.1% | 17 | 20.5% | 16 | 40.0% | 5 | 15.6% | 47 | 39.8% |
| CDR — Clinical Dementia Rating Scale |  |  |  |  |  |  |  |  |  |  |  |  |
| 0=Healthy | - | 0.0% | 19 | 7.0% | 18 | 21.7% | 1 | 2.5% | - | 0.0% | - | 0.0% |
| 0.5=Questionable | 8 | 3.5% | 101 | 37.0% | 62 | 74.7% | 39 | 97.5% | - | 0.0% | - | 0.0% |
| 1=Mild Dementia | 209 | 92.5% | 132 | 48.4% | 3 | 3.6% | - | 0.0% | 32 | 100.0% | 97 | 82.2% |
| 2=Moderate Dementia | 9 | 4.0% | 17 | 6.2% | - | 0.0% | - | 0.0% | - | 0.0% | 17 | 14.4% |
| 3=Severe Dementia | - | 0.0% | 4 | 1.5% | - | 0.0% | - | 0.0% | - | 0.0% | 4 | 3.4% |
| Unknown | 3 | 1.3% | - | 0.0% | - | 0.0% | - | 0.0% | - | 0.0% | - | 0.0% |
| MMSE — Mini-Mental State Examination |  |  |  |  |  |  |  |  |  |  |  |  |
| 0-15 | 4 | 1.8% | 46 | 17.2% | 1 | 1.2% | 1 | 2.5% | 3 | 9.7% | 41 | 35.7% |
| 16-23 | 130 | 59.4% | 153 | 57.3% | 36 | 44.4% | 30 | 75.0% | 20 | 64.5% | 67 | 58.3% |
| 24-30 | 85 | 38.8% | 68 | 25.5% | 44 | 54.3% | 9 | 22.5% | 8 | 25.8% | 7 | 6.1% |
| Unknown | 10 | 4.4% | 6 | 2.2% | 2 | 2.4% | - | 0.0% | 1 | 3.1% | 3 | 2.5% |
| EIC — Equivalent Institutional Care |  |  |  |  |  |  |  |  |  |  |  |  |
| Limited Home Care | 136 | 61.8% | 113 | 43.0% | 53 | 65.4% | 27 | 69.2% | 11 | 37.9% | 22 | 19.3% |
| Adult Home Care | 64 | 29.1% | 116 | 44.1% | 23 | 28.4% | 10 | 25.6% | 18 | 62.1% | 65 | 57.0% |
| Full-Time Care: FTC | 20 | 9.1% | 34 | 12.9% | 5 | 6.2% | 2 | 5.1% | - | 0.0% | 27 | 23.7% |
| Unknown | 9 | 3.9% | 10 | 3.7% | 2 | 2.4% | 1 | 2.5% | 3 | 9.4% | 4 | 3.4% |
| Verbal Outbursts |  |  |  |  |  |  |  |  |  |  |  |  |
| No | 175 | 78.8% | 211 | 80.2% | 69 | 85.2% | 37 | 94.9% | 25 | 86.2% | 80 | 70.2% |
| Yes | 47 | 21.2% | 52 | 19.8% | 12 | 14.8% | 2 | 5.1% | 4 | 13.8% | 34 | 29.8% |
| Unknown | 7 | 3.1% | 10 | 3.7% | 2 | 2.4% | 1 | 2.5% | 3 | 9.4% | 4 | 3.4% |
| Delusions |  |  |  |  |  |  |  |  |  |  |  |  |
| No | 162 | 72.0% | 144 | 54.8% | 58 | 71.6% | 27 | 69.2% | 18 | 62.1% | 41 | 36.0% |
| Yes | 63 | 28.0% | 119 | 45.2% | 23 | 28.4% | 12 | 30.8% | 11 | 37.9% | 73 | 64.0% |
| Unknown | 4 | 1.7% | 10 | 3.7% | 2 | 2.4% | 1 | 2.5% | 3 | 9.4% | 4 | 3.4% |
| Extrapyramidal Signs |  |  |  |  |  |  |  |  |  |  |  |  |
| None/Mild | 187 | 84.6% | 196 | 74.2% | 61 | 76.3% | 28 | 73.7% | 26 | 81.3% | 81 | 71.1% |
| Moderate/Severe | 34 | 15.4% | 68 | 25.8% | 19 | 23.8% | 10 | 26.3% | 6 | 18.8% | 33 | 28.9% |
| Unknown | 8 | 3.5% | 9 | 3.3% | 3 | 3.6% | 2 | 5.0% | - | 0.0% | 4 | 3.4% |
| Needs to be Accompanied when Bathing/Eating |  |  |  |  |  |  |  |  |  |  |  |  |
| No | 200 | 89.7% | 164 | 62.1% | 61 | 75.3% | 29 | 74.4% | 18 | 62.1% | 56 | 48.7% |
| Yes | 23 | 10.3% | 100 | 37.9% | 20 | 24.7% | 10 | 25.6% | 11 | 37.9% | 59 | 51.3% |
| Unknown | 6 | 2.6% | 9 | 3.3% | 2 | 2.4% | 1 | 2.5% | 3 | 9.4% | 3 | 2.5% |
| Needs to be Dressed/Washed/Groomed |  |  |  |  |  |  |  |  |  |  |  |  |
| No | 209 | 94.1% | 170 | 64.6% | 59 | 72.8% | 30 | 76.9% | 21 | 72.4% | 60 | 52.6% |
| Yes | 13 | 5.9% | 93 | 35.4% | 22 | 27.2% | 9 | 23.1% | 8 | 27.6% | 54 | 47.4% |
| Unknown | 7 | 3.1% | 10 | 3.7% | 2 | 2.4% | 1 | 2.5% | 3 | 9.4% | 4 | 3.4% |

The study participants were classified according to initial diagnosis (MCI vs. AD dementia) and their subsequent progression. Among the 273 participants, 123 met criteria for MCI at the initial visit, 83 of whom were consistently diagnosed with MCI (nonconverters) and 40 of whom subsequently converted to AD dementia (converters); 150 met criteria for AD dementia at the initial visit, 118 of whom were consistently diagnosed as AD over follow up (nonreverters) and 32 of whom were subsequently re-diagnosed as non-dementia at a follow up visit (reverters).

The present study was conducted as part of IRB protocol 7258R and approved by the New York State Psychiatric Institute Institutional Review Board; informed consent was obtained from all participants in the study

### Longitudinal Grade of Membership (L-GoM) Model

The L-GoM model was estimated using data from Predictors 2. Its development and internal/external validation was previously described [1, 2]. The key idea behind L-GoM is that the manifestations of a complex disease like AD over patients and over time lead to joint distributions of multiple signs/symptoms with high levels of dependency that can be explained by a small number of latent traits that vary over time for individual patients. Under the L-GoM model, the conditional joint likelihood (same as conditional joint probability) for all observable outcomes can be factorized into a product of conditionally independent terms—one for each combination of patient, time, and clinical-variable outcome—where the conditionality is with respect to the latent traits (the “L-GoM scores”) which are defined to be unobserved random variables. Because we cannot actually measure the relevant latent traits, other approaches are required, and this is why L-GoM is relevant. L-GoM provides a method for estimating the latent traits using the simplest possible set of assumptions at each step of model specification—i.e., using a linear probability model with time-invariant probability-coefficients for the conditional outcome probabilities for each combination of patient, time, and clinical-variable outcome; and treating changes in L-GoM scores as movement along fixed trajectories.

Under the L-GoM model, the clinical status of a subject with AD at any point in time is described by six fixed covariates and 73 time-varying clinical signs and/or symptoms (Table 1; individual items are described in Table S.1 in the Supplementary Materials). All signs and symptoms are coded as dichotomous or polytomous variables, representing the presence/absence of a sign/symptom or the graded severity of a clinical measure (e.g., the different ranges of the MMSE). The L-GoM model summarizes each subject’s current clinical status (i.e., all signs/symptoms measured at a given visit) using a convex combination of four outcome-probability vectors, where the weights for each convex combination are a set of four subject/visit-specific L-GoM scores whose boundary values define the four latent disease subtypes used in the model (Figure 1). In other words, each subject’s clinical status at each visit can be represented using just four numbers—the subject’s L-GoM scores on the four disease subtypes (e.g., 25% subtype 1, 15% subtype 2, 40% subtype 3, and 20% subtype 4). The number four was determined to be the minimum number of subtypes consistent with the conditional likelihood factorization described above [10]. The subtype with the lowest frequency of AD signs/symptoms at initial presentation and the slowest rate of progression was labeled **Subtype 1**. **Subtype 2** has only slightly higher initial frequencies of AD signs/symptoms than subtype 1, but unlike subtype 1, subtype 2 has substantially more rapid rates of progression— more rapid even than subtype 3. **Subtype 3** has higher initial frequencies of behavioral (e.g., verbal outbursts), psychiatric (e.g., delusions), and other signs/symptoms (e.g., depression, agitation) than subtypes 1 and 2, and faster rates of progression than subtype 1. The subtype with the most severe clinical signs/symptoms, and the only one with significant mortality and need for FTC, was labeled **Subtype 4**; all disease progression in the first three subtypes is ultimately toward subtype 4.

**Figure 1:**
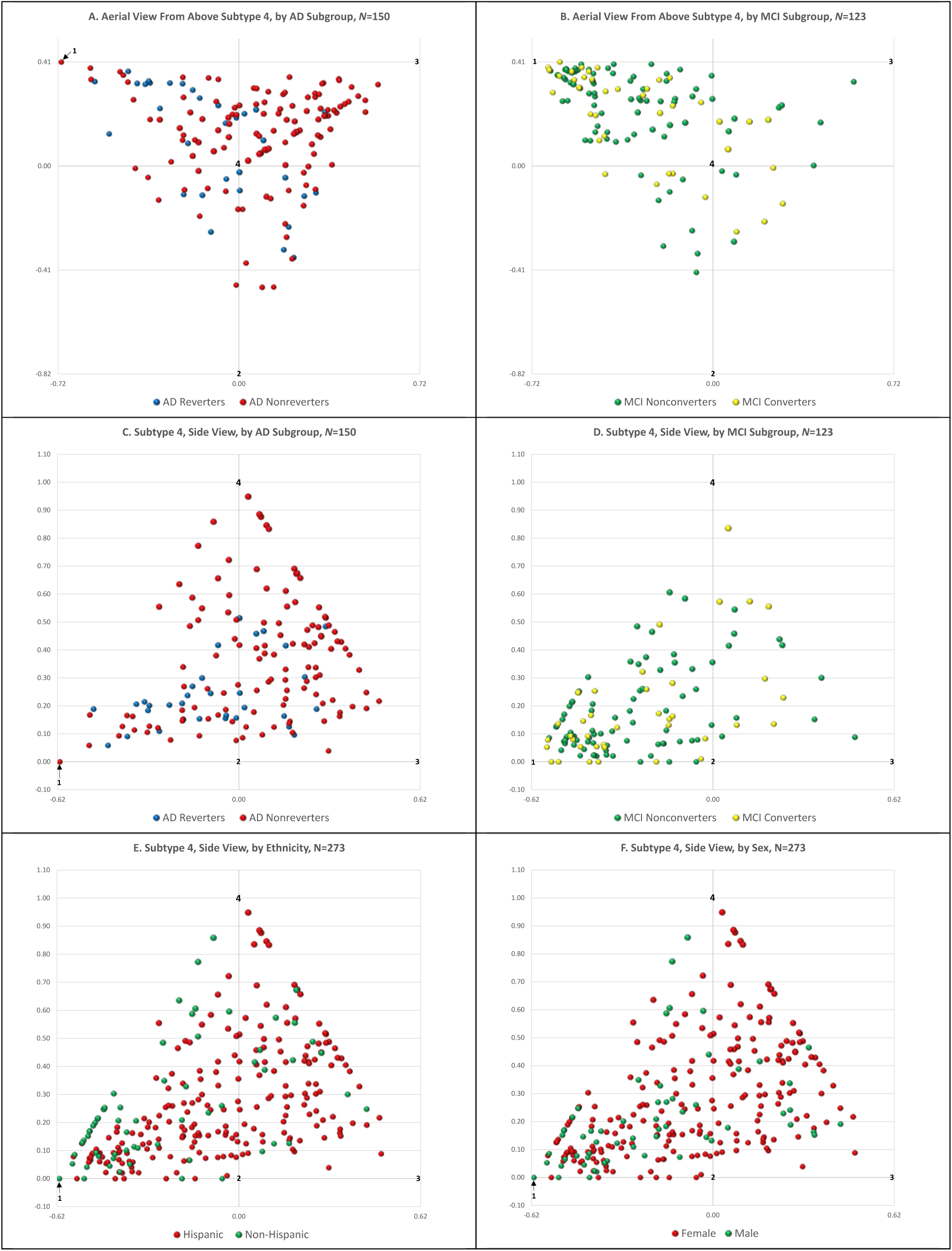
3-D Bubble plot of L-GoM scores at the initial visit for Predictors 3. **Panels A and B**: **Aerial View**. The L-GoM continuum is a regular triangular pyramid (tetrahedron) with a triangular base. Subtypes 1–3 are marked at the vertices of each triangular base, in counterclockwise order. The vertex for subtype 4 is located above the center of the triangular base (i.e., pointing out of the page) where it is also marked. Each bubble represents the estimated L-GoM scores for one subject. The bubbles are color coded by AD subgroup in Panel A and by MCI subgroup in Panel B. The location of each bubble is determined by the subject’s L-GoM scores on subtypes 1–3. The size of each bubble is determined by the subject’s L-GoM score on subtype 4, i.e., its width increases linearly with the subtype 4 score up to a maximum 1.2 times larger at vertex 4 than at the base of the triangular pyramid. The axes in Panels A and B were scaled to the standard GoM metric. **Panels C and D**: **Side View**. These panels show the same L-GoM scores, tetrahedron, and study groups rotated 90 degrees on the subtype 1–3 axis such that the line segment from the center of the triangular base to the vertex for subtype 4 points upward on the page. The *y*-axes in Panels C and D were rescaled to represent the severity of the disease on a scale from 0.0 to 1.0; the same scale factor was also applied to the *x*-axes. With the 90-degree rotation, each triangular base in Panel A and B is contained in its entirety on the *x*-axis, with subtype 4=0.0. The size of each bubble in Panels C and D is unchanged from the size shown in Panels A and B, respectively. The heterogeneity of AD at the initial visit is represented by the distribution of L-GoM scores over the base of the triangular pyramids shown in Panels A and B. By collapsing that distribution to a single point at the centroid of the base, one can generate a 1-dimensional model of AD severity, located at the center of Panels C and D at *x*=0.00, rising vertically. Conversely, by expanding the base of the 1-dimensional model one can reintroduce the heterogeneity on rates of progression (extending along the subtype 1–2 axis) and on behavioral, psychiatric, and other signs/symptoms (extending along the subtype 1–3 axis). **Panels E and F**: **Side View**. These panels show the combined L-GoM scores and triangular pyramid from Panels C and D regrouped by ethnicity and sex, respectively.

The fraction apportioned to each disease subtype for each subject can be any value in the range 0.0–1.0; the sum of the apportionment fractions must be exactly 1.0. The apportionment fractions—or subtype scores—are not directly observed. Instead, they are computationally constructed such that at any point in time a subject’s L-GoM scores reflect, or, alternatively, can be used to generate, the probability of each clinical sign/symptom being present or absent. The constraints on the apportionment ranges and sums imply that the L-GoM scores fall within a regular tetrahedron (equivalently, triangular pyramid). Subtypes 1–3 form the triangular base of the tetrahedron (Figure 1, Panels A and B; more simply, Figs. 1.A and 1.B); subtype 4 defines a perpendicular line segment extending upward from the center of the triangular base (Figs. 1.C–1.F), giving rise to the 3-dimensional structure of the L-GoM model.

The geometry of Figure 1 locates each individual as a point on the boundary or interior of a regular tetrahedron. Each point can be transformed from Euclidian to barycentric coordinates for use in the linear probability model. The barycentric coordinates are the L-GoM scores. This aspect of the model distinguishes it from other latent state models like principal component and factor analysis which do not have a natural transformation from Euclidian to barycentric coordinates, and hence do not support the use of a linear probability model. Further details on the geometry of the model and its relationship to the standard 1-dimensional formulation of AD severity are provided in the legend for Figure 1.

The model describes how a subject’s clinical signs/symptoms progress over the 10-year study period by specifying how the L-GoM scores change between each pair of adjacent visits using a sequence of time-varying upper-triangular transition matrices that are constant across all subjects. This requires us to treat each subject’s L-GoM scores for subtypes 1–4 as an ordered set of four elements—i.e., as a vector—that obeys the rules of matrix algebra. By successively applying the transition matrices to each subject’s vector of L-GoM scores, starting with their initial visit, one can generate their L-GoM vectors at each subsequent visit and, derivatively, the concurrent probability of any clinical sign/symptom included in the model. The transition matrices reported in [1] were smoothed for use in external validation in [2]; this was the only update to the model in the present validation study.

### Mortality and Need for FTC

Two outcomes were used for validation of the L-GoM model predictions in Predictors 3: time of death and need for FTC. Date of death was determined from family report or other sources including the National Death Index, most recently updated through July 2019. For 101 of the 273 subjects, time of death was coded as elapsed time in days since the initial visit. For consistency with Predictors 2, we coded the remaining 172 subjects as surviving to their last visit date with the date/time of death coded as right-censored thereafter.

Need for FTC is part of the Dependence Scale [11]. Using all available information, the examiner ascertained if the care the subject needed and/or received was the equivalent of nursing home care. Of the 273 subjects, FTC information was provided for 263 at the initial visit, at which time 34 needed FTC; FTC information was provided for another 6 subjects only after their initial visit. The subset at risk to needing FTC at each subsequent visit comprised surviving subjects having sufficient information to determine their FTC status.

### Statistical Analysis

Data for each subject’s initial visit were input to the L-GoM model to generate individual-specific L-GoM scores that were subsequently used to generate predicted survival and FTC prevalence curves covering up to 7.5 years beyond the initial visit. The factorization of the conditional joint likelihood for all observable outcomes into a product of conditionally independent terms—one for each combination of patient, time, and clinical-variable outcome—implies that each response is independent of all other responses, conditional on the L-GoM scores. It follows that the conditional joint likelihood of the observed data for each subject does not depend on any missing data for that subject; i.e., the conditional joint likelihood for a given subject involves only the data actually measured. The L-GoM scores for each subject constituted unknown parameters of the conditional joint likelihood for that subject; they were estimated using the maximum likelihood procedures in [1, 2] with all other parameters fixed at the values estimated from Predictors 2. The number of variables used in Predictors 3 was 58; the average number with responses per subject was 54.7, implying that 5.7% of responses were missing (Table 1).

The L-GoM predicted survival curves for individuals were calculated separately and independently at semi-annual intervals following the initial visit for each subject. The L-GoM predicted survival curves for groups/subgroups of subjects defined at the initial visit were calculated by averaging the individual L-GoM predicted survival values at each semi-annual point of time for all members of the respective groups/subgroups. Corresponding observed survival curves were generated for groups/subgroups of subjects using the Kaplan-Meier (KM) product-limit estimator [12] with: (1) mortality recorded at the end of the semi-annual interval during which each death occurred; and (2) censoring/withdrawal without a known date of death recorded at the time of the last completed visit. Simultaneous 95% confidence bands were calculated and centered on the L-GoM predicted survival probabilities using the normal approximation for binomial proportions, with sample sizes and *z*-scores computed as follows. The sample sizes of the respective groups/subgroups at the initial visit were downwardly adjusted at follow-up visits, as needed, to reflect the cumulative effects over time of censoring/withdrawal using eqn. (2j) in [12]. The *z*-score of 1.96 in the conventional normal approximation for binomial proportions was replaced with an adjusted *z*-score of 3.16 so that the 95% confidence bands centered on the L-GoM survival probabilities were approximately the same size as Nair’s simultaneous “equal precision” (EP) 95% confidence bands for the KM estimator—with parameters *a* = 1 – *b* = 0.05; see Table 2 in [13].

The L-GoM predicted FTC prevalence curves for individuals were calculated separately and independently for all subjects with Dependence Scale information at the initial visit and for all surviving subjects with Dependence Scale information at any of the seven annual follow-up visits. The L-GoM predicted FTC prevalence curves for groups/subgroups of subjects were calculated by averaging the individual L-GoM predicted FTC prevalence probabilities at each visit for all members of the respective groups/subgroups with the requisite Dependence Scale information. Corresponding observed FTC prevalence curves were generated for groups/subgroups of subjects treating the observed relative frequencies of need for FTC at baseline or among survivors to each follow-up visit with complete Dependence Scale assessments as binomial proportions. Simultaneous 95% confidence bands were calculated and centered on the L-GoM predicted FTC prevalence probabilities using the normal approximation for binomial proportions, with the *z*-score of 1.96 in the conventional normal approximation for binomial proportions replaced with an adjusted *z*-score of 2.73, using Bonferroni’s method to determine the adjusted *z*-score needed to control for the eight simultaneous comparisons [14].

The confidence bands were centered on the predicted survival and predicted prevalence curves, with the width of the confidence interval at each predicted plot point taking account of the number of subjects supporting the corresponding observed plot points. With this centering, the probability that at least one observed plot point falls outside the resulting 95% confidence bands is approximately 5%—under the null hypothesis that the L-GoM model is the correct model. Rejection of the null hypothesis is equivalent to rejection of the L-GoM model. Ideally, for validation, one would want all observed plot points within the 95% confidence bands, in which case one would accept (i.e., “fail to reject”) the L-GoM model. Practically, one could provisionally accept the model if only one or two plot points were barely outside the 95% confidence bands, i.e., if the deviations were reasonably small [15]. In order to explore the impact of lack of fit for AD reverters, the survival and FTC prevalence analyses were replicated after excluding the AD reverters.

Between-group comparisons of the L-GoM scores at the initial visit were conducted separately, but not independently, for each of the four subtypes via two-tailed Student’s *t*-tests, using Bonferroni’s method (i.e., multiplying each raw *p*-value by 4) to adjust the *p*-values for each *t*-test for multiple-testing at the overall 5% level of significance—equivalent to, but more convenient than, increasing the conventional large-sample critical value for each *t*-test from 1.96 to 2.50 [16]. The overall null hypothesis—that the average initial L-GoM scores do not differ between the groups—was rejected if the adjusted *p*-value was less than 0.05 for any *t*-statistic. The logic is similar to that of the *M*-test for the multinomial distribution [17]. In each case, a single summation constraint (i.e., L-GoM scores and multinomial probabilities must each sum to 1.0) leads to dependencies between the component test statistics that were controlled using Bonferroni’s method.

### Statistical Software

Statistical calculations were performed using SAS 9.4, Excel 2016, Simply Fortran 2.41, and R 3.5.1.

## RESULTS

### Demographics

Demographic characteristics of the Predictors 2 and 3 cohorts are displayed in Table 2 along with baseline values for eight time-varying variables selected from the 11 domains listed in Table1. The characteristics for Predictors 3 are also displayed separately for MCI nonconverters (*N*=83) and converters (*N*=40), and AD reverters (*N*=32) and nonreverters (*N*=118). Compared to Predictors 2, the Predictors 3 cohort was relatively older, more female, and more Hispanic, with lower educational attainment and lower frequency of the ApoE e4 allele (APOE4^+^).

### L-GoM Scores and Subtypes

Figure 1 displays the L-GoM scores for all subjects at the initial visit in Predictors 3, grouped as in Table 2, with supplementary displays by sex and ethnicity. Table 3 compares the average L-GoM scores for the groups in Figure 1, with supplementary comparisons for ApoE. The average L-GoM scores differed significantly between the MCI and AD groups, with the average on subtype 1 higher for MCI and the average on subtypes 3 and 4 higher for AD. The average L-GoM scores differed significantly between AD reverters and AD nonreverters, with the average on subtype 1 higher for reverters and the average on subtype 4 higher for nonreverters. The average L-GoM scores did not differ significantly between MCI converters and MCI nonconverters.

**Table 3.** Average L-GoM Scores by Group/Subgroup at Initial Visit, Predictors 3.

|  | Group/Subgroup |  | *t*-statistic | Adjusted *p*-value |
| --- | --- | --- | --- | --- |
| Diagnosis | MCI | AD |  |  |
| N/N/df/Overall *p* | 123 | 150 | 271 | <0.001 |
| Subtype 1 | 0.567 | 0.241 | 10.63 | <0.001 |
| Subtype 2 | 0.104 | 0.126 | -1.07 | NS |
| Subtype 3 | 0.150 | 0.297 | -7.05 | <0.001 |
| Subtype 4 | 0.180 | 0.337 | -6.85 | <0.001 |
| MCI | Nonconverter | Converter |  |  |
| N/N/df/Overall *p* | 83 | 40 | 121 | NS |
| Subtype 1 | 0.573 | 0.556 | 0.33 | NS |
| Subtype 2 | 0.099 | 0.112 | -0.42 | NS |
| Subtype 3 | 0.151 | 0.147 | 0.11 | NS |
| Subtype 4 | 0.177 | 0.185 | -0.23 | NS |
| AD | Reverter | Nonreverter |  |  |
| N/N/df/Overall *p* | 32 | 118 | 148 | <0.01 |
| Subtype 1 | 0.364 | 0.207 | 3.38 | <0.01 |
| Subtype 2 | 0.174 | 0.113 | 1.76 | NS |
| Subtype 3 | 0.225 | 0.317 | -2.46 | NS |
| Subtype 4 | 0.237 | 0.364 | -3.17 | <0.01 |
| Sex | Male | Female |  |  |
| N/N/df/Overall *p* | 60 | 213 | 271 | <0.001 |
| Subtype 1 | 0.525 | 0.349 | 4.13 | <0.001 |
| Subtype 2 | 0.072 | 0.128 | -2.30 | NS |
| Subtype 3 | 0.191 | 0.242 | -1.87 | NS |
| Subtype 4 | 0.212 | 0.281 | -2.32 | NS |
| Ethnicity | Non-Hispanic | Hispanic |  |  |
| N/N/df/Overall *p* | 62 | 211 | 271 | <0.001 |
| Subtype 1 | 0.507 | 0.353 | 3.62 | <0.01 |
| Subtype 2 | 0.106 | 0.119 | -0.51 | NS |
| Subtype 3 | 0.128 | 0.261 | -5.14 | <0.001 |
| Subtype 4 | 0.259 | 0.268 | -0.28 | NS |
| ApoE | APOE4<sup>-</sup> | APOE4<sup>+</sup> |  |  |
| N/N/df/Overall *p* | 188 | 85 | 271 | <0.01 |
| Subtype 1 | 0.417 | 0.323 | 2.43 | NS |
| Subtype 2 | 0.113 | 0.122 | -0.41 | NS |
| Subtype 3 | 0.205 | 0.286 | -3.38 | <0.01 |
| Subtype 4 | 0.265 | 0.269 | -0.16 | NS |

Notes: *p*-values adjusted for four simultaneous *t*-tests using Bonferroni's method; each overall *p* is the most significant of the corresponding adjusted *p*-values; NS denotes nonsignificance (i.e., adjusted *p* ≥ 0.05). See text for details.

The average L-GoM scores differed significantly by sex, with the average on subtype 1 higher for males; by ethnicity, with the average on subtype 1 higher for non-Hispanics and the average on subtype 3 higher for Hispanics; and by ApoE, with the average on subtype 3 higher for APOE4^+^. Among AD nonreverters (not shown): (1) the average on subtype 1 was higher for males (*t*=3.42, adj. *p*<0.01); (2) the average on subtype 3 was higher for Hispanics (*t*=3.24, adj. *p*<0.01); and (3) the average on subtype 3 was higher for APOE4^+^ (*t*=3.67, adj. *p*<0.01).

### Mortality

Figure 2 presents the observed and predicted survival curves both overall (Fig. 2.A) and for five subgroups (Figs. 2.B–2.F). Figure 2.A shows that the model-based curves provided close fits to the observed data with just one observation (at 54 months) clearly outside the simultaneous confidence bands and a second observation (at 6 months) overlaying but just outside its confidence interval. Given that the 54-month deviation was small relative to the confidence band and the observed points following that deviation were within the confidence bands at all later visits, this fit may be good enough to use the model in out-of-sample applications.

**Figure 2:**
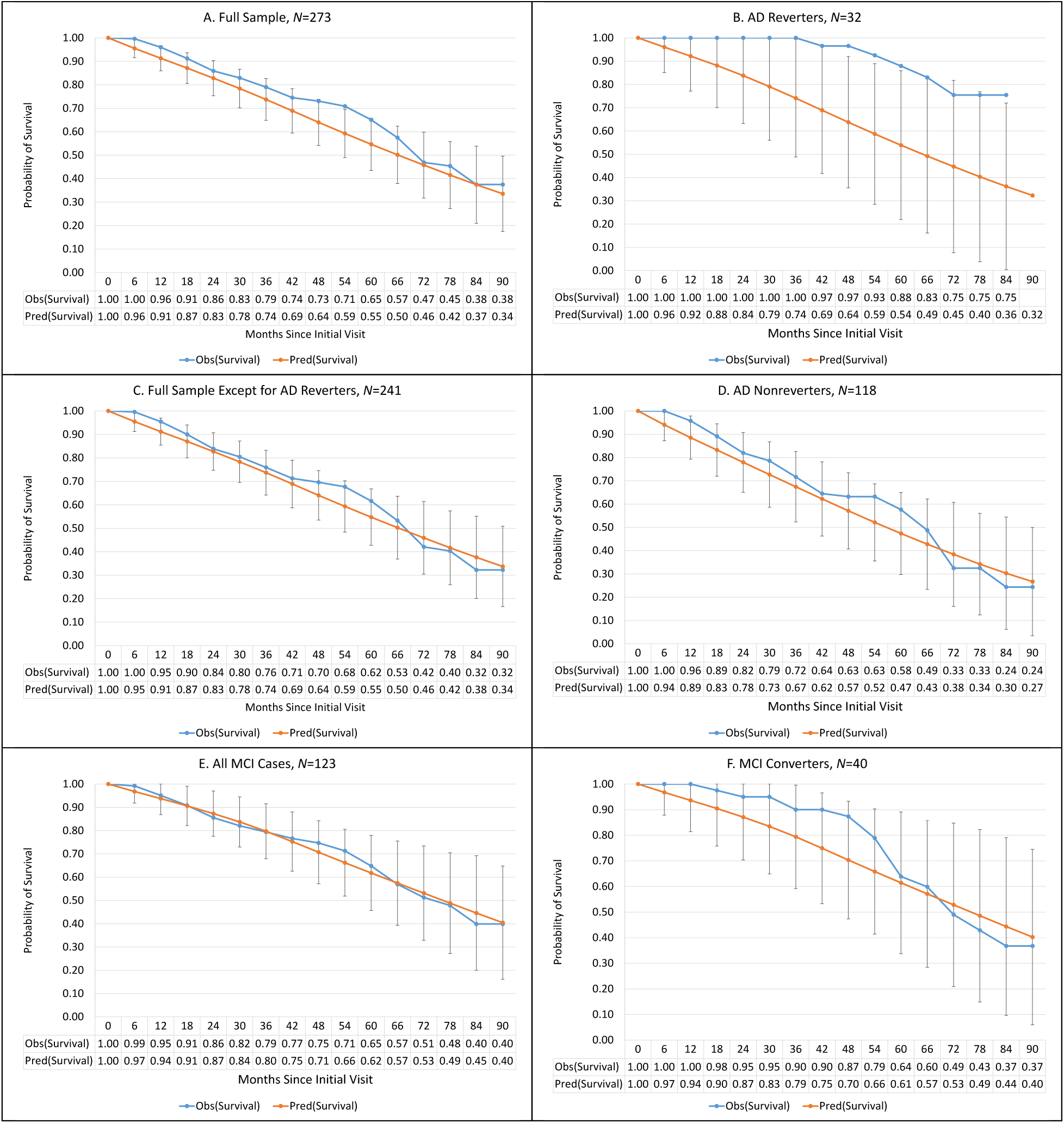
Observed vs. predicted probability of survival in Predictors 3 under L-GoM model derived from Predictors 2, with 95% simultaneous confidence intervals based on Nair’s “Equal Precision” bands (Nair, 1984) (with parameters *a* = 1 – *b* = 0.05). Curves for average survival are shown in the upper left panel (A) for *N* = 273 subjects where *N* = Number of subjects seen at the initial visit. The *x*-axis indicates the number of months since the initial visit. Corresponding survival curves are shown in the remaining panels for subsets of the study sample defined using: (B) AD reverters; (C) all subjects except AD reverters; (D) all AD nonreverters; (E) all MCI cases; and (F) all MCI converters.

Nonetheless, Predictors 3 has several well-defined subgroups that adequately explain the deviations in Figure 2.A. The observed survival for the 32 AD reverters was substantially higher than predicted (Fig. 2.B) and higher than observed for the rest of the Predictors 3 cohort (Fig. 2.C). Only five of the 32 AD reverters died during follow-up, and none during the first 36 months; seven observations in Figure 2.B were outside the confidence bands; four were visibly noticeable. The model does not fit these data.

Figure 2.C replotted the data in Figure 2.A excluding the 32 AD reverters. All observations in Figure 2.C were within the confidence bands; hence, the model fits these data. Figure 2.D displays the results for the 118 AD nonreverters. All observations in Figure 2.D were within the confidence bands; thus, the model fits these data. Figure 2.E displays the results for the 123 subjects with MCI; the fit was excellent. Figure 2.F shows that the 40 MCI converters survived longer than predicted, though not long enough to fall outside the confidence bands. A complementary shortening effect was manifest for the 83 MCI nonconverters (not shown)— sufficient, on balance, to produce the very close fit shown in Figure 2.E. Hence, Figure 2.E validates the L-GoM model for MCI.

The median predicted survival among MCI cases was 19.7 months above that of the AD nonreverters (76.4 vs. 56.7 months, respectively; Figs. 2.E and 2.D). The combined median predicted survival for these two groups was 66.4 months (Fig. 2.C).

Figure 3 stratifies Figure 2.C by ethnicity and sex. All observed values in Figure 3 were within the associated confidence bands except for Hispanics and females, at 12 months, where the deviations were less than the thickness of the plot point. The predicted survival curves for Hispanics and non-Hispanics were similar—with the Hispanic median lower by 1.1 month— consistent with their initial higher scores on subtype 3. The predicted survival curve for males was higher than for females—with the male median higher by 11.5 months—consistent with their initial higher scores on subtype 1.

**Figure 3:**
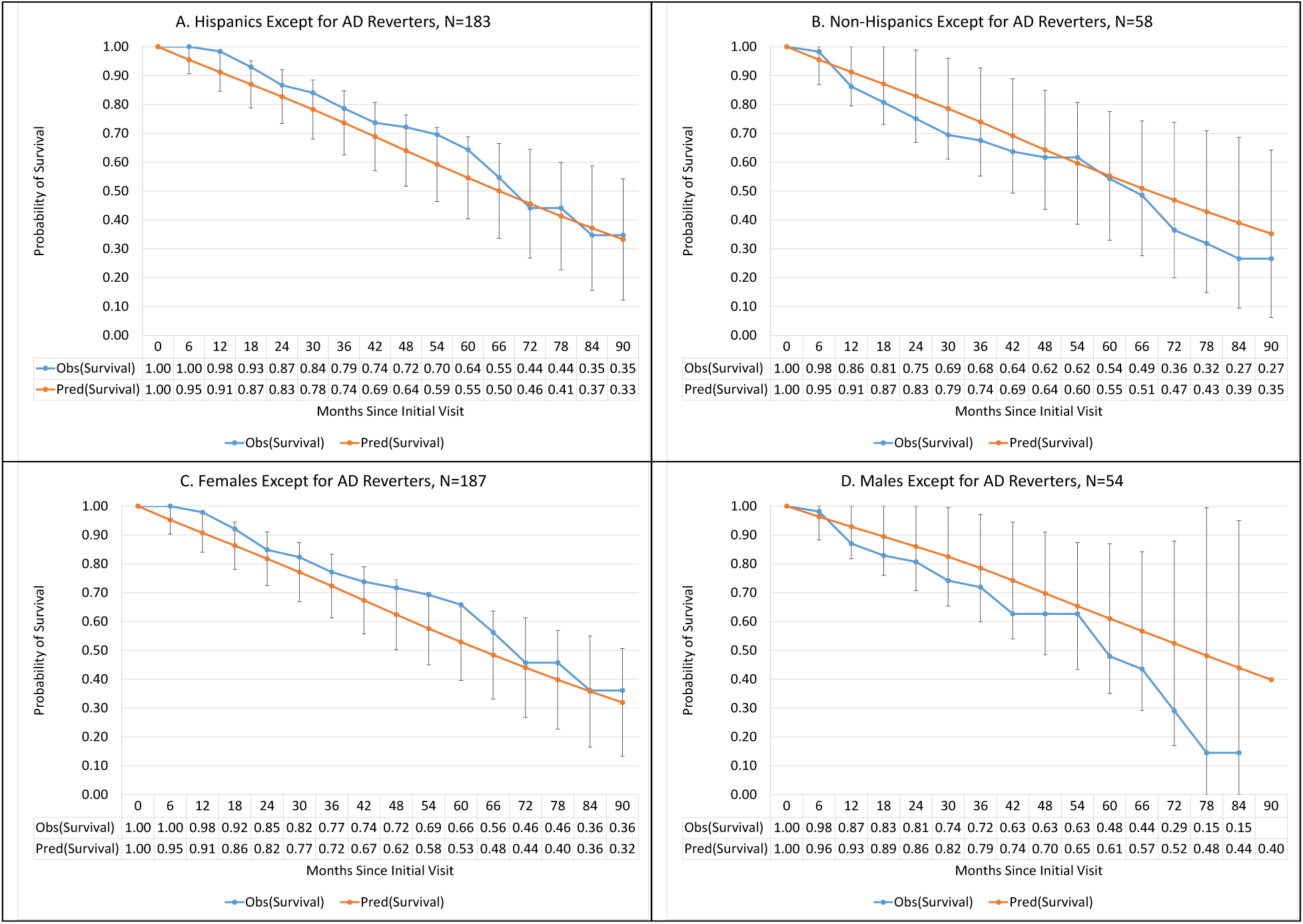
Observed vs. predicted probability of survival in Predictors 3, for all subjects except AD reverters, under L-GoM model derived from Predictors 2, with 95% simultaneous confidence intervals based on Nair’s “Equal Precision” bands (Nair, 1984) (with parameters *a* = 1 – *b* = 0.05). Curves for average survival are shown in the upper left panel (A) for *N* = 183 Hispanic subjects where *N* = Number of subjects seen at the initial visit. The *x*-axis indicates the number of months since the initial visit. Corresponding survival curves are shown in the remaining panels for subsets of the study sample defined using: (B) non-Hispanics; (C) females; and (D) males.

### Need for FTC

Figure 4 displays the prevalence curves for need for FTC, both overall (Fig. 4.A) and for the same five subgroups (Figs. 4.B–4.F) shown in Figure 2. The title line of each panel indicates the number (*N*) of subjects contributing one or more visits in which FTC status was assessed and the total number of visits (*V*) contributed by the *N* subjects. The confidence bands are wider than in Figure 2 because the prevalences in Figure 4 are conditional probabilities for which the number of subjects at risk decreases at least as fast as the survival curves in Figure 2; it decreases further if information on FTC status is missing for subjects at the respective visit.

**Figure 4:**
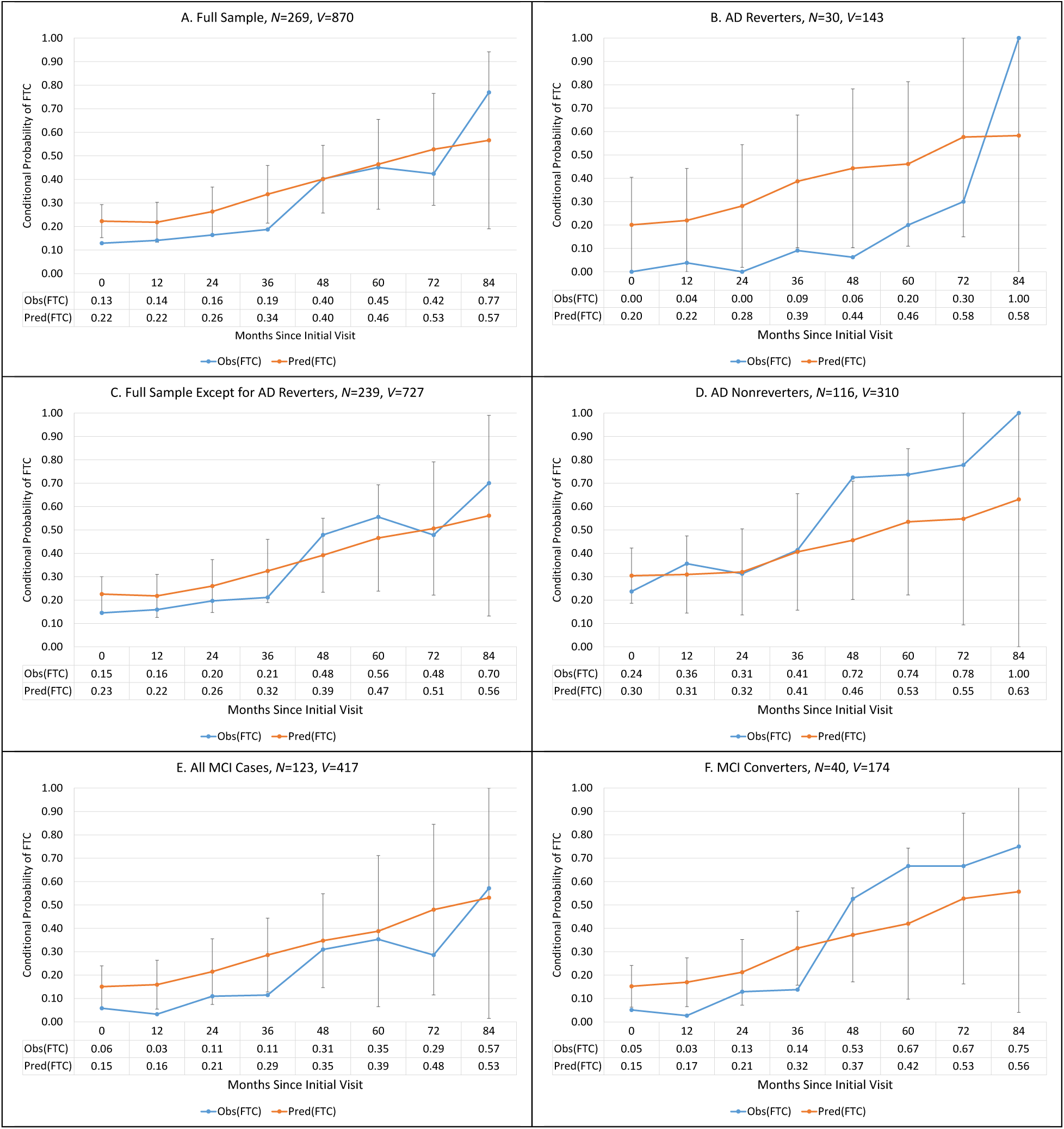
Observed vs. predicted conditional probability of need for FTC among survivors to each follow-up visit in Predictors 3 with complete Dependence Scale assessments under L-GoM model derived from Predictors 2, with 95% simultaneous confidence intervals using Bonferroni’s correction for multiple testing (Dunn, 1961). Curves for overall average prevalence of need for FTC are shown in the upper left panel (A) for *N* = 269 subjects with *V* = 870 visits where *N* = Number of subjects contributing at least one visit and *V* = Total number of Visits contributed by the *N* subjects. The *x*-axis indicates the number of months since the initial visit. Corresponding prevalence curves are shown in the remaining panels for sample subsets using: (B) all AD reverters in Panel A; (C) all subjects in Panel A except AD reverters; (D) all AD nonreverters in Panel A; (E) all MCI cases; and (F) all MCI converters.

The model-based curves in Figure 4.A overpredicted the observed curves in the first 36 months; two observations were outside the confidence bands—at 0 and 36 months. The observed points converged to and crossed over the model predictions following the second deviation.

Figure 4.B shows that the observed probabilities of FTC for 30 (of 32) AD reverters were substantially lower than for Figure 4.A up to 84 months, consistent with their better survival (Fig. 2.B); three observations (at 24, 36, and 48 months) were outside the confidence bands.

Figure 4.C replotted Figure 4.A excluding the 30 AD reverters. All observations but the first in Figure 4.C were within the confidence bands; the first deviation was less than the thickness of the plot point. Thus, the model in Figure 4.C was close to fitting the data.

Figure 4.D displays the results for 116 (of 118) AD nonreverters. All observations except at 48 months were within the confidence bands; the deviation at 48 months was relatively small. The observed and predicted curves were almost identical from 0 to 36 months—the predictions were excellent for this time period. The observed prevalences beyond 48 months were higher than predicted but still within the confidence bands.

Figure 4.E displays the results for all 123 MCI cases; there were two visits (at 12 and 36 months) where the observed prevalence rates were visibly below the confidence bands and a third observation (at 0 months) overlaying but just below its confidence band. The observed and predicted prevalence rates agreed closely beyond 36 months.

Figure 4.F shows that the 40 MCI converters had lower than predicted FTC prevalence rates up to 36 months and higher than predicted thereafter; all observed rates thereafter were within the confidence bands. The observed prevalence rates for the 83 MCI nonconverters (not shown) were uniformly lower than predicted, contributing to the deviations in Figure 4.E.

Figure 5 stratifies Figure 4.C by ethnicity and sex. All observed values in Figure 5 were within the associated confidence bands except for Hispanics at 0 months where the deviation was slightly greater than the thickness of the plot point. The predicted prevalence curves for Hispanics and non-Hispanics were similar up to 36 months despite initial differences on subtype 3. The predicted prevalence curve for males was substantially lower than for females at all visits—consistent with their initial higher scores on subtype 1.

**Figure 5:**
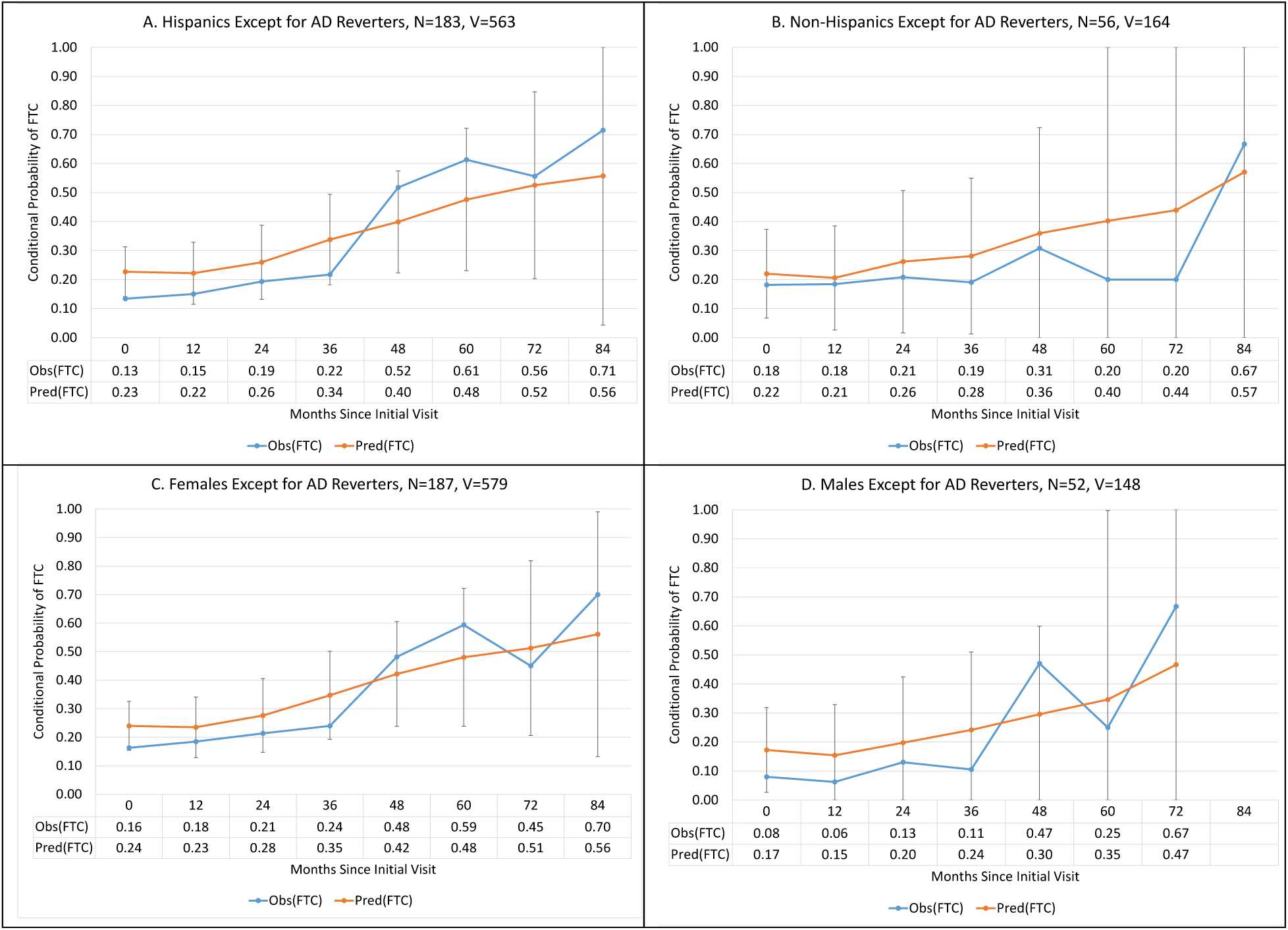
Observed vs. predicted conditional probability of need for FTC among survivors to each follow-up visit in Predictors 3 with complete Dependence Scale assessments, for all subjects except AD reverters, under L-GoM model derived from Predictors 2, with 95% simultaneous confidence intervals using Bonferroni’s correction for multiple testing (Dunn, 1961). Curves for overall average prevalence of need for FTC are shown in the upper left panel (A) for *N* = 183 Hispanic subjects with *V* = 563 visits where *N* = Number of subjects contributing at least one visit and *V* = Total number of Visits contributed by the *N* subjects. The *x*-axis indicates the number of months since the initial visit. Corresponding prevalence curves are shown in the remaining panels for sample subsets using: (B) non-Hispanics; (C) females; and (D) males.

### Individual Survival Functions—Calibration

The L-GoM scores in the present study were estimated using the same code as in our web-based calculator described in the Supplementary Materials. Thus, the L-GoM validation in the present study also serves to validate the web-based calculator.

To gain insight into the accuracy of the model, we present in Figure 6 the observed and predicted survival curves for four distinct L-GoM subgroups (numbered 1–4). The subgroups were formed from the 241 subjects in Figure 2.C by determining which, if any, of the corresponding L-GoM scores on subtypes 1–4 were > 0.50. This can occur just once per subject, if at all, due to the constraint that the sum of the L-GoM scores = 1.00. The subgroups contained *N* = 91, 10, 28, and 36 subjects for subtypes 1 to 4, respectively, with 76 subjects ungrouped. The median predicted survival for subgroups 1–4 were 86.0, 59.9, 66.5, and 37.3 months, respectively, and 59.7 months for the ungrouped subjects (not shown). All observations in Figure 6 were within the subgroup-specific confidence bands; thus, the model fits the data. The medians for the subgroups can be compared with the overall median predicted survival of 66.4 months for the 241 subjects (Fig. 2.C). The 48.7-month spread between the medians for subgroups 1 and 4 was 2.5 times larger than the 19.7-month spread between AD nonreverters and MCI cases (Figs. 2.D and 2.E). The individual medians ranged from 25.4 months (lowest in subgroup 4) to 108.2 months (highest in subgroup 1)—a high-low ratio of 4.3 to 1— demonstrating that the L-GoM scores can be highly informative predictors for individuals with AD or MCI.

**Figure 6:**
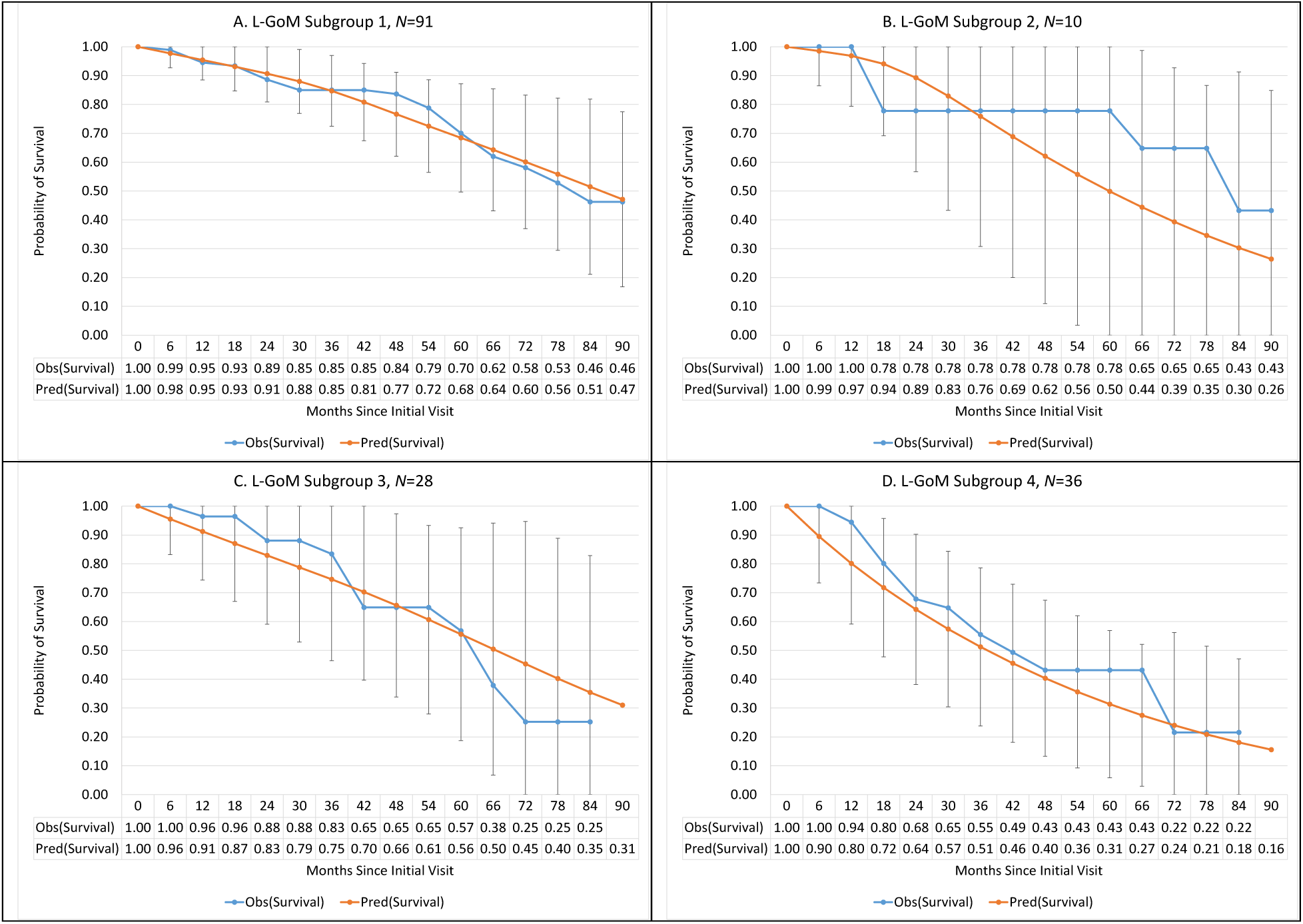
Observed vs. predicted probability of survival in Predictors 3, excluding AD reverters, under L-GoM model derived from Predictors 2, with 95% simultaneous confidence intervals based on Nair’s “Equal Precision” bands (Nair, 1984) (with parameters *a* = 1 – *b* = 0.05). Panel (A): Survival curves for subgroup 1 with *N* = 91 subjects having subtype 1 score > 0.50. The *x*-axis indicates the number of months since the initial visit. Corresponding survival curves are shown in the remaining panels for: (B) subgroup 2, comprising *N* = 10 subjects having subtype 2 score > 0.50; (C) subgroup 3, comprising *N* = 28 subjects having subtype 3 score > 0.50; and (D) subgroup 4, comprising *N* = 36 subjects having subtype 4 score > 0.50.

The use of the Nair [13] bands in Figure 6 followed the same logic as in Figures 2–3 with one important difference—the subgroups were selected from the extremes of the distribution of the L-GoM scores (Figure 1) to uncover any deficiencies in the model. None were found. Thus, the model was well-calibrated for the overall sample, for males, females, Hispanics, non-Hispanics, MCI, and AD nonreverters, and for the four extreme L-GoM subgroups. Importantly, the model was well-calibrated for the entirety of the 90-month follow-up since the initial visit.

### Individual Survival Functions—Discrimination

Validation on extreme subgroups is a common approach for determining if a model can be confidently used for individuals with unusually low or unusually high mortality risks [18]. Another approach for determining this aspect of the model uses the concordance index [19, 20] to compare observed vs. predicted survival times under the model. For the 241 subjects considered above, the concordance index was *C* = 0.610 (95%-CI 0.546–0.674), indicating that, of two randomly chosen subjects, the subject with the higher predicted life expectancy had a 61% chance of outliving the subject with the lower predicted life expectancy. Note that *C* = 0.610 is roughly equivalent to *R*^2^ = 0.114 [21].

A third approach uses a time-specific AUC statistic to compare the predicted survival function values at a given time *t* between subjects who died at time *t* (the “cases”) and those who survived beyond time *t* (the “controls”). Heagerty and Zheng [22] showed that the concordance index is a weighted average of the time-specific AUC statistics, implying that the aforementioned 0.610 concordance index provides an overall summary of the time-specific AUC statistics for our data.

A fourth approach redefines the time-specific AUC statistic to include all deaths at or prior to time *t* among the “cases.” This “cumulative” approach is used when scientific interest focuses on discrimination at a specific time *t*, or among a small collection of such times [22]. Based on Figure 6, the range 36–60 months was of prime interest in the present study because it contained the median survival times for subgroups 2 and 4, as well as for the ungrouped subjects. The cumulative AUC statistics were 0.660 (36 mo.; 95%-CI 0.570–0.750), 0.680 (42 mo.; 95%-CI 0.596–0.764), 0.684 (48 mo.; 95%-CI; 0.601–0.767), 0.657 (54 mo.; 95%-CI 0.572–0.742), and 0.590 (60 mo.; 95%-CI 0.497–0.682), with all but the last significantly higher than 0.50; the 95%-CIs were computed using eqn. (1) in [23].

A fifth approach combines aspects of the first four. It starts by validating the calibration of the individual survival functions, both overall and for extreme subgroups, as under the first approach. Its focus then shifts to quantifying and visually displaying the calibration and discrimination properties of the model simultaneously, building on the histograms of the hazard ratios derived from the L-GoM survival function values for the 241 subjects considered above; the results are illustrated in Figure 7 for *t*=36 and *t*=54 months since initial visit—at which times the observed cumulative AUC statistics were 0.660 and 0.657, respectively. Each set of hazard ratios was scaled to have a mean of 1.0; the associated variances were 0.369 for 36-month exposure (Fig. 7.A) and 0.249 for 54-month exposure (Fig. 7.B). To smooth the histograms, we fitted separate gamma distributions to the hazard ratios by matching moments and confirmed that the fits were statistically acceptable (*p*>0.05 each, using one-sample Kolmogorov-Smirnoff tests).

**Figure 7:**
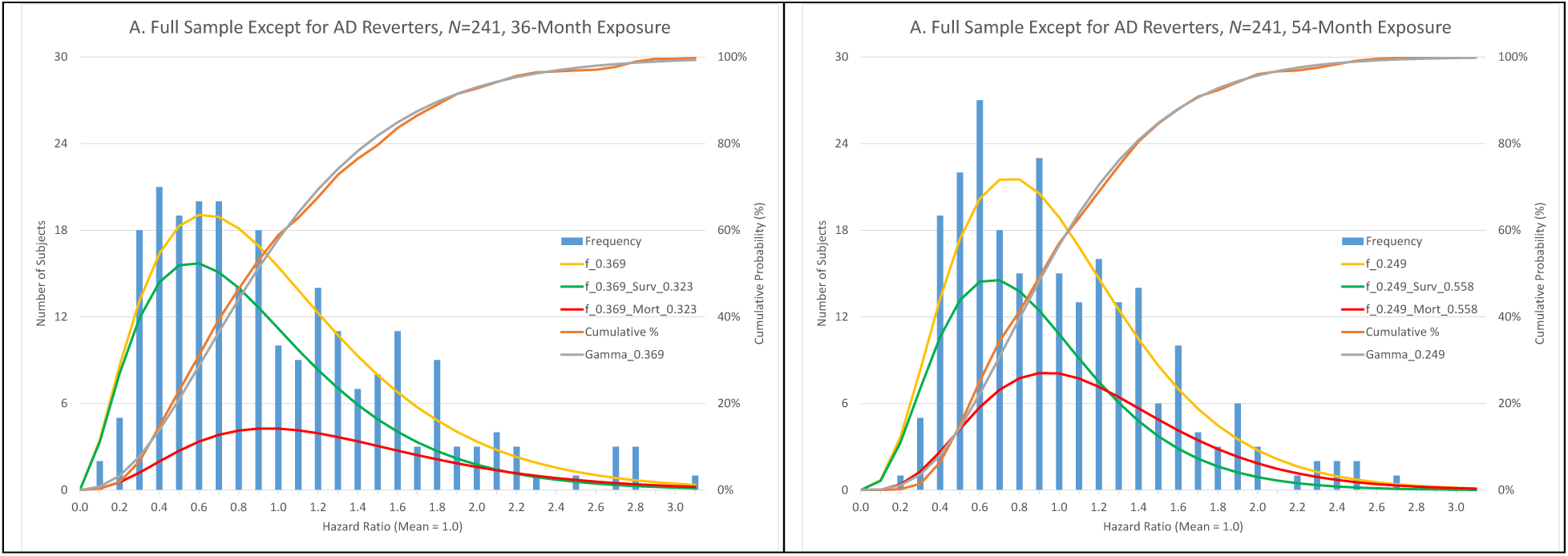
Histograms of the 36- and 54-month hazard ratios (displayed separately in Panels A and B) from the L-GoM model for the 241 subjects shown in Figure 1.C overlaid with separately fitted gamma distributions. The left vertical axes indicate the number of subjects (ranges 0–22 and 0–27) represented by the blue bars in each histogram. The right vertical axes indicate the level (range 0–100%) of the observed (orange line) and fitted (gray line) cumulative distribution functions. The bottom horizontal axes represent the 36- and 54-month hazard ratios (ranges 0.1–3.1 and 0.2–2.7), computed as follows. Panel A—The individual-specific 36-month cumulative hazards were obtained from the natural logarithms of the individual-specific survival function values at 36 months since initial visit (see Figures 2, 3, and 6). Because the natural logarithms were negative, we changed their signs before continuing. The average over the 241 subjects of the individual-specific 36-month cumulative hazards defined the mean 36-month cumulative hazard (0.323). The hazard ratios were formed using the individual-specific cumulative hazards as the numerators and the mean cumulative hazard as the denominator. Thus, the hazard ratios had a mean of 1.0, by design. The variance of the hazard ratios was 0.369. Panel B—Same as Panel A except that the exposure time was 54 months (i.e., 50% longer), the mean cumulative hazard was 0.558, and the variance of the hazard ratios was 0.249. The correlation of the hazard ratios in Panels A and B was 0.994. We fitted gamma density functions (gold lines in Panels A and B) to the hazard ratios by matching moments, with each density function scaled to the sample size (241). We assessed the goodness of fit of the gamma cumulative distributions (orange lines) to the observed cumulative distributions (gray lines) using one-sample Kolmogorov-Smirnoff tests (*p*>0.05 each; the fits were acceptable). The fitted density functions (gold lines) indicated that the hazard ratios were unimodal and right-skewed. The fitted density functions were decomposed into two complementary sub-distribution density functions corresponding to survivors (green lines) and decedents (red lines), respectively, at 36 and 54 months since initial visit. The hazard ratios at each tick mark on the horizontal axis in Panel A were multiplicatively paired with the mean 36-month cumulative hazard (0.323) to compute the survival and mortality probabilities for that tick mark; for Panel B, the corresponding pairing was done with the mean 54-month cumulative hazard (0.558). The sub-distributions for the decedents correspond to the “cases” in the 36- and 54-month cumulative AUC statistics while the sub-distributions for the survivors correspond to the “controls”.

Following the logic of the cumulative AUC statistic, each fitted density function (gold lines) was decomposed into two additive sub-distributions: one for survivors (“controls”; green lines) and the other for decedents (“cases”; red lines) over the indicated exposure time. The decomposition clarified how the observed cumulative AUC statistics (0.660 and 0.657, respectively) related to the prognostic hazard ratios: there was about a 66% probability that a randomly selected decedent had a higher hazard ratio than a randomly selected survivor. The model-based cumulative AUCs (0.690 and 0.673) closely matched the observed cumulative AUCs—indicating that the model-based cumulative AUCs were well-calibrated. The mean hazard ratio for decedents was 1.45 times larger than for survivors at 36 months and 1.34 times larger at 54 months.

The case-control logic of the cumulative AUC statistics allows the hazard ratios to be regressed on survival status, effectively reversing causality. Applying this logic to the sub-distributions in Figs. 7.A and 7.B (using [24]), we obtained *R*^2^ values of 0.086 and 0.087, respectively. To aid interpretation, we considered counterfactuals where the hazard ratios were exponentially distributed (i.e., with means and variances set to 1.0; retaining the same average survival probabilities), yielding “improved” *R*^2^ values of 0.194 and 0.241, with the respective mean hazard ratios for decedents 2.36 and 2.67 times larger than for survivors. The maximum *R*^2^ value (0.348) occurred for the counterfactual to Fig. 7.A where the variance of the gamma distribution was set to 4.1 (i.e., with the same mean, but even more right-skewed than the exponential), for which the mean hazard ratio for decedents was 10.5 times larger than for survivors. We concluded that it would be difficult to increase our *R*^2^ values absent a substantially more right-skewed distribution of hazard ratios; alternatively, our *R*^2^ values can be interpreted using the *R*^2^ values from the exponential distribution as a benchmark.

It follows that there will remain substantial irreducible variation in the individual survival times after accounting for the individual-specific hazard ratios with the individual survival curves exhibiting patterns similar to those shown in Figure 6.

## DISCUSSION

The three Predictors Studies share common instruments which makes Predictors 3 ideal for external validation of our latest implementation of the L-GoM model.[1][2] The original version of L-GoM was developed using Predictors 1 and validated on preliminary data from Predictors 2 [10, 25]. Following the completion of data collection for Predictors 2, we determined that the improved design (e.g., including ApoE, MMSE, and full BDRS) and the later fielding of Predictors 2 would work better for a re-specified version of L-GoM using pooled male/female data to increase sample size and facilitate identification of sex differences [1]. The new version of L-GoM had separate treatment of fixed vs. time-varying covariates and was optimized for predictive applications using data from the first visit alone. This version was validated using Predictors 1 [2]. The success of the Predictors 1 validation was not surprising, however, given the previous reverse-order validation in [10].

Predictors 3 differed from Predictors 2 in design, recruitment, and target population [3]. Moreover, Predictors 3 included three subgroups minimally represented in Predictors 2—MCI converters, MCI nonconverters, and AD reverters. These subgroups offered the opportunity to expand the scope of application of the L-GoM model. The distributional differences with respect to race, ethnicity, and education offered further opportunities to expand the scope of application of the L-GoM model beyond the clinic-based high SES non-Hispanic white patients enrolled at specialized AD centers with mild AD dementia at their initial visit in Predictors 1 and 2. The WHICAP-based recruitment protocol ensured that Predictors 3 participants were representative of MCI and AD dementia cases in North Manhattan with respect to sex, age, race, ethnicity, and education—making the present study generalizable to the local area population, and possibly beyond to other urban areas. The high percentage of Hispanic participants in Predictors 3 allowed the present study to assess AD progression in this important but understudied segment of the U.S. population noted for its high incidence and prevalence of AD dementia [26, 27].

The target population for the L-GoM model validation was older persons with probable AD dementia. This target was represented in the Predictors 3 Study by the 118 AD nonreverters. The results in Figures 2.D and 4.D provided strong support for the validity of the L-GoM model for this target sample. These results contrasted sharply with those for the 32 AD reverters— supporting the assumption that these cases were incorrectly diagnosed as having AD at the initial visit, given that a major hallmark of AD is its progressive nature. Identification of possible misdiagnosis of AD dementia based on reversion to lower CDR scores required information not available at the initial visit. Almost all (i.e., 29 of 32) reversions occurred within 2–3 years after the initial visit and almost all (i.e., 28 of 32) were for the Hispanic group. Nonetheless, 12 of the 32 AD reverters (11 Hispanics) had a second reversal in which they exhibited a CDR score of 1 or 2 at a later visit—suggesting that their AD diagnoses may have been premature rather than incorrect. Applying the same reversion rules to Predictors 2, we identified seven (of 229) potential AD reverters of whom six exhibited a CDR score of 1 or 2 at a later visit.

The higher prevalence of reverters in Predictors 3 than in Predictors 2 most likely relates to Predictors 2 (and Predictors 1) being clinic-based while Predictors 3 was community-based. Patients seek clinic-based diagnoses when cognitive, functional, or behavioral problems are severe enough to raise concerns that enhance the probability that the diagnosis of AD will be accurate. In contrast, participants in Predictors 3 were chosen at random from the community. In a community-based study where the participants did not self-select to seek a diagnosis, it is more likely that they meet diagnostic criteria due to causes unrelated to AD, thereby accounting for the higher prevalence of AD reverters in Predictors 3.

### L-GoM Scores and Subtypes

The goal of L-GoM was to infer from the longitudinal data the latent traits underlying the clinical progression of AD. As noted above, the conditional likelihood factorization combined with the linear probability model implied that the latent traits can be represented as points in the L-GoM continuum—in our case a tetrahedron. It follows that the longitudinal changes in the latent traits can be represented as movement within the L-GoM continuum. The Predictors 2 visits were 6 months apart, which allowed up to 21 such points to be generated for each participant over the 10-year follow-up (with fewer points for those who died during follow-up).

The trajectories account for increases in severity over time; different subjects follow different trajectories. This latent structure accounts for heterogeneity in initial presentation and in rates of disease progression. To facilitate prediction using only the information from visit 1, we assumed that once the initial point in the L-GoM continuum was determined, then all future points along the trajectory were fixed—a key simplifying assumption of L-GoM.

The substantially higher initial average score on subtype 4 for AD nonreverters in Predictors 3 (0.364) than in Predictors 2 (0.099) implies substantially greater AD severity at the initial visit. In addition to the 23.7% of AD nonreverters in need of FTC at the initial visit, another 57.0% needed adult home care vs. 9.1% and 29.1%, respectively, in Predictors 2. Similarly, 17.8% of AD nonreverters had CDR>1 vs. 4.0% in Predictors 2; 35.7% of AD nonreverters had MMSE<16 vs. 1.8% in Predictors 2; and 55.1% of AD nonreverters were 85+ years vs. 14.0% in Predictors 2.

The initial average score on subtype 4 for the MCI cases in Predictors 3 was also substantially higher than for the AD cases in Predictors 2—attributable, in part, to their older ages (56.1% aged 85+ vs. 14.0% in Predictors 2), higher prevalence of moderate/severe extrapyramidal signs (24.6% vs. 15.4%), greater need to be accompanied when bathing/eating (25.0% vs. 10.3%), and greater need to be dressed/washed/groomed (25.8% vs. 5.9%). More than offsetting this difference, however, the average subtype 1 score for the MCI cases in Predictors 3 (0.567) was substantially higher than for the AD cases in Predictors 2 (0.291), consistent with MCI having lower initial frequencies of multiple items from the Dependence Scale and the Blessed Dementia Rating Scale [1] (Table 2).

### Mortality

The survival predictions in Figures 2.C–2.E support the validity of the L-GoM model in Predictors 3. In contrast, the survival predictions in Figure 2.B identify an important subgroup in Predictors 3—AD reverters—for which the L-GoM model was not valid. How should we interpret this finding? The positive results in Figures 2.C–2.E combine with the finding that all subjects in Figure 2.B reverted to CDR scores below the dementia threshold to suggest that this subgroup did not have AD dementia. Most likely, the majority also did not have MCI and were not at-risk for AD.

The accuracy of the survival predictions for participants with MCI (Figure 2.E) supports the validity of the extension of the model to MCI. This extension meets a published goal in conducting the Predictors 3 Study [3]—made possible because the L-GoM model’s four subtypes characterize a heterogeneous range of AD presentations and disease severities at the initial visit. With subtype 1 having the lowest frequency of AD signs/symptoms, the 123 MCI cases had an average initial score on subtype 1 of 0.567 vs. an average of 0.207 for the 118 AD nonreverters. Conversely, with subtype 4 representing the L-GoM model’s disease severity scale, the 123 MCI cases had an average initial score on subtype 4 of 0.180 vs. an average of 0.364 for the 118 AD nonreverters.

One further aspect of Figure 2 was the absence of observed mortality in the first six months. This may be an artifact of measurement in community-based epidemiologic studies in which individuals more likely to die within six months are less likely to agree to participate. The effect was short-term and not impactful beyond 6–12 months.

### Need for FTC

The predictions of need for FTC in Figures 4.C–4.E further support the validity of the L-GoM model in this population. Unlike the survival predictions in Figure 2, the predictions in Figure 4 were for conditional probabilities of need for FTC at the indicated visits. Another difference is the need for FTC was based on judgments of trained interviewers applying the Dependence Scale protocol whereas mortality was unequivocal. Figure 4.D indicates that the FTC predictions for 116 AD nonreverters were excellent for 0–36 months but possibly a little low beyond that time. Figure 4.E indicates that the FTC predictions for 123 MCI cases were possibly a little high up to 48 months after which they appear to be very good. Figure 4.C, which combined these two groups, indicates an acceptable overall fit.

Figures 4.C–4.E have important implications for analyses that treat need for FTC as a patient outcome or endpoint of an AD cohort study (e.g., see [28] and [1]). Figure 4.D shows that the predicted prevalence of FTC was above 30% at the initial visit for AD nonreverters. While this was above the observed initial prevalence in Predictors 3, the order reversed at 12 months when the observed prevalence was above 35%. This presents challenges to the use of FTC as an endpoint of treatment/intervention with no obvious way to represent the impact of the initial FTC group in, for example, a standard Cox [29] regression model. Moreover, 24 of the 110 subjects who needed FTC at one visit had a later visit at which they did not need FTC, indicating that FTC is not a stable endpoint. L-GoM analysis resolves both issues: it allows FTC to be analyzed without special accommodations for non-zero initial FTC levels or for subsequent reversals in FTC levels.

### Combining FTC with Survival

The L-GoM model provides a way to predict the probability of future need for FTC at the time of a patient’s initial visit: one needs only to multiply the predicted conditional FTC probability at the selected future time (e.g., Figures 4–5) by the predicted probability of surviving to that time (e.g., Figure 2–3, 6). This method facilitates prediction of total future time in need of FTC during the remaining lifetime of individual patients with new diagnoses of MCI or AD dementia; see [1]. Our web-based calculator uses these calculations to compute the predicted residual years of life to be lived with/without need for FTC.

### Calibration and Discrimination

Calibration refers to the accuracy of a model’s predictions across a range of relevant outcome probabilities both overall and for specified subgroups (e.g., Figures 2–6). Good calibration for extreme subgroups means that the model can be used prognostically for subjects with unusually low or unusually high risks (e.g., Figure 6).

Discrimination refers to the ability of a model to separate subjects with good outcomes from those with poor outcomes. The assessment of the discriminative power of any model of AD survival is inherently difficult because there are no “good” outcomes. Death is certain; only the timing is uncertain. This differs fundamentally from other applications of the concordance index [19] or AUC statistic [22], where good clinical outcomes are possible (e.g., successful treatment, disease remission, etc.). The presentation of AD at the initial visit is heterogeneous; the L-GoM model allows this heterogeneity to be quantified by a set of hazard ratios that summarize all information available at the initial visit with respect to survival over any fixed follow-up exposure time *t*. When the hazard ratios at the initial visit are scaled to have a mean of 1.0 then the discriminative power of the model at time *t* can be summarized in two statistics: the variance of the hazard ratios and the mean cumulative hazard up to time *t*. If one holds the mean cumulative hazard up to time *t* fixed, then the larger the variance, the greater will be the separation of the hazard ratios for decedents vs. survivors. Similarly, if one holds the variance of the hazard ratios fixed, then the larger the mean cumulative hazard up to time *t*, the greater will be the separation of the hazard ratios for decedents vs. survivors. If one decreases the variance but increases the mean cumulative hazard up to time *t*, then the separation of the hazard ratios may be relatively stable, as demonstrated in Figure 7. If the model is well-calibrated, then the predicted cumulative AUC statistics will closely match the observed cumulative AUC statistics, as also demonstrated in Figure 7.

### Generalizability

With each successive validation study, our goal has been to extend the scope of applicability of the L-GoM model. The Predictors 3 Study was essential to this goal in that the participants constituted a random sample of Medicare enrollees in a single large multiethnic urban community in North Manhattan [3]. We know from the experience of the Framingham Heart Study that there is much to be learned from such community studies, despite the obvious need for further generalization. Our prior estimation/validation studies on Predictors 1 and 2 used clinic-based racially/ethnically homogeneous cohorts, with unknown generalizability, which was resolved using Predictors 3.

The three Predictors Studies were designed to allow development of predictors of disease course among persons with AD. The L-GoM model was specifically designed to focus on predictors of severity in AD. The input variables are listed in Table 1. The predictors of severity are the L-GoM scores shown in Figure 1, which are readily generated for current and new subjects using our web-based calculator. The impact of the L-GoM scores on survival can be summarized in the form of mortality hazard ratios, as shown in Figure 7. The L-GoM hazard ratios change gradually over time as the length of the follow-up period is extended. For example, the hazard ratios in Figure 7.A based on the 36-month cumulative hazard functions were correlated 0.991–0.999 with similarly constructed hazard ratios using the 24–30 and 42– 60-month cumulative hazard functions.

Our results strongly supported the conclusion that the empirical hazard ratios were unimodally distributed and right-skewed for the 36- and 54-month exposure times. The discriminative power was sufficiently high for the model to be used for individual-level prognostication if users are cautioned against treating the median survival predictions as predictions of actual survival time.

Our results were broadly consistent with two recent studies. First, Schaffert et al. [30] applied stepwise multiple regression to data from the National Alzheimer’s Coordinating Center (NACC) to analyze elapsed time to death following the visit at which AD diagnosis occurred among 764 autopsy-confirmed AD cases with an overall mean time to death of 53.4 months. Their best model yielded an overall *R*^2^ value of 0.274 with seven predictors (sex, age, race/ethnicity, MMSE, neuropsychiatric symptoms, abnormal neurological exam, and functional impairment) obtained from an initial set of 21 predictors; MMSE was the strongest predictor, accounting for 0.204 of the overall 0.274 *R*^2^. The overall *R*^2^ value was interpreted to mean that users should not treat the regression predictions as predictions of actual survival time. Stratified analyses for persons with mild, moderate, and severe MMSE (21–30, 13–20, and 0–12, respectively) at the initial visit yielded *R*^2^ values of 0.056, 0.115, and 0.159, respectively, with mean times to death of 67.1, 54.5, and 35.8 months. The 31.3-month difference between the mean times to death for mild vs. severe MMSE was broadly consistent with the 48.7-month difference between the medians for subgroups 1 and 4 in our model (Figure 6). The stratified *R*^2^ values were consistent with the analysis of hazard ratios for decedents vs. survivors in our model (Figure 7).

Second, Deardorff et al. [31] used survival data from 4,267 participants in the Health and Retirement Study (HRS) with probable dementia to generate a Cox proportional hazards regression model with 12 predictors, including four types of comorbidities (cancer, heart disease, diabetes, and lung disease) but excluding tests of cognitive ability comparable to the MMSE—Schaffert’s [30] strongest predictor. Deardorff et al. [31] reported good calibration and good discrimination with time-specific AUCs of 0.73 (95%-CI 0.70–0.76) at 1 year, 0.72 (95%-CI 0.70–0.75) at 2 years, and 0.74 (95%-CI 0.71–0.76) at 5 years in validation data from the National Health and Aging Trends Study (NHATS) (N=2,404). Their AUCs were 0.04–0.08 higher than our best AUCs; our 95%-CIs overlapped theirs. The interquartile ranges on their individual-specific median predictions (their Figure 2.B) were roughly comparable to the interquartile ranges in our Figure 6. Unlike Schaffert et al. [30], they did not include direct measures of dementia severity among their predictors.

Deardorff’s [31] eFigure 5 indicated that the baseline hazard ratios in NHATS were more skewed than in Predictors 3 (cf. Figure 7). To better understand this difference, we compared the upper tails of the distribution of predicted probabilities of death during the first two years after baseline in each study. Deardorff’s [31] Figure 1 grouped the 2-year predicted probabilities by decile, with the decile-specific average probabilities for deciles 8–10 being 0.37, 0.45, and 0.61, respectively (vs. 0.40, 0.46, and 0.59 observed). The corresponding average 2-year predicted probability for decile 10 in Predictors 3 was 0.38 (vs. 0.41 observed), implying that decile 10 in Predictors 3 most closely matched decile 8 in NHATS, but with no matches for deciles 9 and 10—suggesting that the case-inclusion protocols used by Deardorff et al. [31] were more successful than the case-recruitment protocols in Predictors 3 in identifying subjects with dementia who had unusually high mortality risks. Deardorff [31] noted that such high mortality risks may be due to unrelated comorbid diseases; e.g., a subject randomly selected from decile 10 had 4 ADL dependencies, 5 IADL difficulties, cancer, lung disease, and diabetes at the baseline visit.

Wolfson et al. [32] cautioned that epidemiological studies routinely and unintentionally exclude substantial fractions of dementia cases with unusually high mortality risks. Such exclusion is an inherent feature of the sampling design whenever the rapid progression of the disease itself prevents inclusion of affected cases in the study. The effect on summary statistics can be substantial; e.g., Wolfson et al. [32] reported that the observed median survival time of 6.6 years among 821 dementia cases in the Canadian Study of Health and Aging decreased to 3.3 years after adjustment for such exclusion. On the other hand, the effect on individual-specific estimates can be minor. Indeed, Wolfson’s [32] adjustment for the summary statistics was based on the assumption that the individual-specific estimates were correct. An important implication is that the individual-specific prognoses derived from such epidemiological studies can be substantially more accurate than the unadjusted average prognoses from the same studies.

### Limitations

Our prior studies [1, 2] showed that the L-GoM model applies to non-Hispanic whites— comprising 90.5% and 90.2% of the Predictors 1 and 2 cohorts, respectively. The present study shows that the L-GoM model applies to Hispanics—comprising 77.5% of the overall Predictors 3 cohort and 82.9% of AD nonreverters. The Hispanics in Predictors 3, however, were predominantly (90.5%) of Caribbean origin; e.g., 72.5% were Dominican. The present study also shows that the L-GoM model applies to non-Hispanic nonwhites—predominantly (94.6%) African Americans—but the sample size (*N*=37) was too small to be fully confident in the validation for these populations. Our validation now needs to be replicated for other minority groups in the United States.

The present study shows that misdiagnosis of AD (evidenced by reversion to lower CDR scores) can lead to incorrect predictions; similarly, though not detected in the present study, misdiagnosis of MCI can also yield incorrect predictions. Misdiagnosis of AD was predominantly (87.5%) for Hispanics; non-Hispanic nonwhites accounted for the remaining 12.5% with no AD misdiagnoses among non-Hispanic whites.

The L-GoM model employs a 3-dimensional structure with four component subtypes (Figure 1) [1] derived from 11 measurement domains (Table 1) using L-GoM-score estimation procedures described in [2]. The model can accommodate other sources of heterogeneity by expanding one or more of the existing vertexes from a single point to a new axis extending to additional subtypes. The current and prior validation studies indicate, however, that such expansion is unnecessary; i.e., the current 3-dimensional structure is valid for modeling AD presentation and progression for the three Predictors Studies. Subsequent application of the L-GoM model to other longitudinal studies may require modification to accommodate differences in instrumentation for the 11 existing measurement domains and biological domains not currently represented, including AD imaging, CSF biomarkers, postmortem pathologic data, and differential gene expression profiles. Further research is needed to determine if other biological domains can be accommodated within the existing 3-dimensional structure—potentially linking the current set of clinical signs/symptoms to the underlying pathology in a unified model of AD progression.

The L-GoM model was designed to accurately represent the clinical progression of AD over the 10-year period following the initial assessment/diagnostic visit for AD. All variables, including death and need for FTC, were given equal priority in estimation. Unrelated comorbidities were not included in our model. In contrast, the models developed by Schaffert et al. [30] and Deardorff et al. [31] were specifically optimized to predict survival/mortality among persons with autopsy-confirmed AD or probable dementia. Deardorff et al. [31] emphasized that unrelated comorbidities can be important predictors in such models. When such comorbidities are manifest, their separate impact on survival should be considered, e.g., using Deardorff’s [31] calculator.

Our validation procedure emphasized the use of simultaneous confidence bands based on Nair’s [13] or Bonferroni’s [14] method of construction. The advantage of simultaneous confidence bands over pointwise confidence bands was that the simultaneous bands provided a formal method of testing the goodness of fit over the entirety of the 90-month follow-up time since the initial visit. Pointwise bands provide only an informal method of evaluation and are difficult to interpret when a small number of observed points are just outside the bands. For example, with just 20 points plotted in a given figure, one would expect that, on average, one observed point would lie outside the pointwise confidence band—implying that such bands provide no basis to reject the fit of the model. The graphical displays in Figures 2–6 allow readers to judge for themselves the goodness of fit of the model and, combined with Figure 7, its suitability for use in their applications.

### Conclusions

The major aims of the three Predictors Studies were to further our understanding of AD progression in order to develop algorithms to predict the length of time from disease onset to major disease outcomes in individual patients with AD. Our results showed that the initial presentation of AD is quite heterogeneous, incorporating signs/symptoms from one or more of 11 domains (Table 1). The initial heterogeneity across the 11 domains can be represented as an ordered set of four L-GoM scores (Figure 1) with subtype 4 representing the model’s disease severity scale, i.e., subtype 4 was the only subtype with significant mortality and need for FTC. Our validation showed that mortality and need for FTC could be accurately predicted for all subjects except AD reverters—who were likely misdiagnosed as having AD. We concluded that the L-GoM model can be used to predict outcomes in community-dwelling older adults of different ethnic, linguistic, and SES backgrounds. In addition, we found for the first time that the model extends to MCI.

## SUPPLEMENTARY MATERIALS

The web-based calculator implementing the L-GoM model calculations is described in the Supplementary Materials.

## Supporting information

Supplement: AD Risk Calculator

## Data Availability

All data produced in the present study are available upon reasonable request to the authors

## Acknowledgments

Research reported in this publication was supported by the National Institute on Aging of the National Institutes of Health under Award Number R01AG007370. The content is solely the responsibility of the authors and does not necessarily represent the official views of the National Institutes of Health.

## Conflicts Of Interest

Dr. Stern consults for Eisai, Lilly, and Arcadia. Columbia University licenses the Dependence Scale, and in accordance with university policy, Dr. Stern is entitled to royalties through this license. The remaining authors report no conflicts of interest.

## Notes

### Author Declarations

The present study was conducted as part of IRB protocol 7258R and approved by the New York State Psychiatric Institute Institutional Review Board.

### Summary of Updates

Revised text with additional figure Supplement with more extended description of ris calculator

