## Supplement: AD Risk Calculator for "Validation of a Multivariate Prediction Model of the Clinical Progression of Alzheimer’s Disease in a Community-Dwelling Multiethnic Cohort"

This Supplement provides a brief description of the AD Risk Calculator developed in association with the main article and provides the Web-Link needed to access the current version of the calculator. It provides a guide to the items utilized by the calculator and discusses interpretations and limitations of the calculator’s outputs.

**Brief Description**

The calculator allows the user to input available information without requiring the user to guess answers to questions not assessed by an individual’s health care provider; in other words, missing data does not constitute a problem for the calculator. After data are entered, pressing the “Run Calculator” button produces the predictive information described below. At that point, adding additional data will modify the information.

The calculator provides individual-specific survival probabilities extending from the present time throughout the next 10 years. The calculator uses demographic and other fixed information in combination with time-varying disease-relevant information covering 11 domains of measurement generally available from the initial baseline assessment of individuals diagnosed with mild cognitive impairment (MCI) or Alzheimer’s disease (AD) dementia. The variables for the specific inputs to the calculator are listed in Table S.1.

The calculator also provides individual-specific probabilities related to the need for full-time care (FTC) extending from the present time throughout the next 10-year period, using the same baseline information as for the survival probabilities. The definition of FTC is equivalent to the level of care provided in a nursing home, but it also includes such care provided at home or in community-based assisted living facilities.

The calculations employ the longitudinal Grade of Membership (L-GoM) model to compute the probability of mortality over each successive 6-month interval as well as the probability of needing FTC at each 6-month anniversary of the initial assessment. An in-depth explanation of the methodology can be found in the *Alzheimer's Research & Therapy* article cited below in References.

**Web-Link**

The Web-Link to the calculator is: <https://cnd-columbia.shinyapps.io/AD_Calc/>

**Instructions**

The user should answer all questions where the answer is known with reasonable certainty. There is no requirement that all questions be answered. Unanswered questions are treated as "Not Applicable." As more questions are answered, the accuracy of the estimated survival and FTC probabilities improves. A suggested minimum includes Demographics, Function, Dependence, Psychiatric/Depression issues, and Mini Mental State evaluation score (if available).

Answers to one or more questions can be changed following the initial "Run Calculator" command; the survival and FTC probability estimates will be re-calculated accordingly.

**Interpretation**

Alzheimer’s disease exhibits substantial variability in its initial presentation and rate of progression. The variability in rate of progression is strongly related to the variability in the specific signs and symptoms identified at the initial baseline presentation. The calculator uses the initial signs and symptoms for a given individual to guide it in calculating the probabilities of survival and of the onset of need for FTC. It calculates the probability of each outcome at 6-month intervals over the next 10 years associated with the given individual’s values on the signs and symptoms at the baseline assessment. These calculations are summarized in two graphical displays—the survival curve and the FTC curve—described in the next several paragraphs.

The survival curve plots the probability of being alive at each 6-month anniversary of the baseline assessment with interpolated values for times between the 6-month plot points. The survival curve starts at the value 1.0—because the probability of being alive at time 0 is exactly 100%—and decreases gradually over the next 10 years towards the value 0.0. The time at which the survival curve reaches the value 0.50 is the median survival time—i.e., the 50^th^ percentile of the distribution of survival times. The summary table below the survival curve reports survival times at the 90^th^, 75^th^, 50^th^, 25^th^, and 10^th^ percentiles.

The FTC curve plots the joint probability of being alive and free of the need for FTC at each 6-month anniversary of the baseline assessment with interpolated values for times between the 6-month plot points. The FTC curve generally begins at a value below 1.0 at time 0, reflecting the empirical fact that many individuals are in need of FTC at the time of their initial assessment. The FTC curve lies below the survival curve at all times beyond the baseline assessment—because it is a joint probability curve—and it decreases faster than the survival curve—due to increases in the need for FTC over time.

The FTC curve is interpreted by reference to the survival curve at the same time. The FTC curve separates each survival value into two additive components: (1) a component free of need for FTC; and (2) a component needing FTC. The first value can be read off the *y*-axis; the second value can be obtained by subtracting the first value from the associated survival value. The summary table below the FTC curve reports the FTC components at the 90^th^, 75^th^, 50^th^, 25^th^, and 10^th^ percentiles of the distribution of survival times.

The FTC curve also separates the entire area below the survival curve into two distinct regions: (1) a region free of need for FTC; and (2) a region needing FTC. The relative sizes of each region represent the fractions of life during the next 10-year period free of need for FTC vs. needing FTC.

**Limitations**

The calculator **does not** provide predictions of the actual length of survival or actual onset of need for FTC for any individual. Instead, the calculator uses the baseline assessments to generate summary scores that can best predict the rate of progression of the disease for each individual. Individuals with identical summary scores will have identical predictions of their rates of progression. In this way, it is technically correct to consider a small group of individuals having identical summary scores. Of 100 such individuals observed at time 0, the calculator predicts that 50 would die prior to the median survival time and 50 would die later. The calculator, however, does not predict which specific individuals would die before the median versus after the median; that prediction **cannot** be done. Similar considerations apply to the interpretation of the FTC curves.

Despite such limitations, knowing their median survival times is still of great value to individuals with AD, their caregivers, and health care providers. Based on the results in the main article, we determined that the individual-specific median survival times range from 25 to 108 months depending on the initial signs and symptoms. Comparing these values with the overall median of 66 months in the study sample, one can conclude that the survival probability for individuals can be very different from the averages for the populations to which they belong. Similarly, the fractions of life during the next 10-year period lived in need of FTC range from 13% to 79% with an overall median of 37%. Again, the range of variation is quite substantial.

**Acknowledgments**

Based on the algorithm described in the *Alzheimer's Research & Therapy* article cited above, the prototype for the AD Risk Calculator was funded as a Pilot Project to Duke University under grant number P30AG034424 from the National Institute on Aging of the National Institutes of Health, with PIs Eric Stallard and Kenneth C. Land; the calculator was developed by Joseph Lucas and Courtney Page. Research supporting estimation of the parametric inputs to the calculator was funded at Columbia University by grant number R01AG007370 from the National Institute on Aging of the National Institutes of Health, with PI Yaakov Stern; this grant funded subsequent modifications and upgrades to the calculator made at Duke University by David Straley and at Columbia University by Seonjoo Lee and Hyunnam Ryu. The content of the AD Risk Calculator is solely the responsibility of the authors and does not necessarily represent the official views of the National Institutes of Health.

**Table S.1: AD Risk Calculator Inputs**

This table summarizes the individual items that can be entered into the calculator. If a value is unknown, then it can be left blank; not all values need to be filled in. In the main article, data for each of the values was based on the instrument noted in the table. Full names for instrument abbreviations and references for the instruments can be found below the table. Many of the items can be entered without access to the specific instrument. Prompts in the input portion of the program provide information about the question that is asked and the range of possible responses. However, some inputs require specific scores, such as the scores on items from the Mini-Mental State examination.

| *Variable* | *Description* | *Source* |
| --- | --- | --- |
| APOE | APOE genotype | Personal report |
| Gender | (Male, Female) | Personal report |
| Age at Intake | Current age | Personal report |
| Race | (White, Non-White) | Personal report |
| Occupation |  | Personal report |
| Years Since Diagnosis |  | Personal report |
| Adequate Sight? | Can the patient see adequately? | Personal report |
| Adequate Hearing? | Can the patient hear adequately? | Personal report |
| Beer/Week | Beer: # of 12 oz. bottles/week | Personal report |
| Wine/Week | Wine: # of 14 oz. glasses/week | Personal report |
| Hard liquor/Week | Hard liquor: # of 1 oz. jiggers/week | Personal report |
| Living Status | Where does the patient live currently? | Personal report |
| Years since entered LTC Facility | If the patient lives in a long-term care facility other than home, how long has the patient stayed there? | Personal report |
| Wandered Away | Has the patient wandered away from home or from the caregiver? (past month) | CUSPAD |
| Verbal Outbursts | Has the patient made verbal outbursts? (past month) | CUSPAD |
| Physical Threats | Has the patient used physical threats and/or violence? (past month) | CUSPAD |
| How Much Sleep? | Amount of sleep (past month) | CUSPAD |
| Patient Trouble With Chores | Inability to perform household tasks | BDRS |
| Patient Trouble Handling Money | Inability to cope with small amount of money (past year) | BDRS |
| Patient Trouble Remembering Lists | Inability to remember shortlist of items; for example, in shopping list (past year) | BDRS |
| Patient Trouble Around House | Inability to find way about indoors (past year) | BDRS |
| Patient Trouble Around Neighborhood | Inability to find way about familiar street (past year) | BDRS |
| Patient Trouble Recognizing Place | Inability to interpret surroundings (past year) | BDRS |
| Patient Trouble Remembering Things | Inability to recall recent events (past year) | BDRS |
| Patient Dwell in the Past | Tendency to dwell in the past (past year) | BDRS |
| Patient Eating | Problems with eating independently (past year) | BDRS |
| Patient Dressing | Problems with dressing independently (past year) | BDRS |
| Patient Bladder and Bowel Control | Problems with using the bathroom independently (past year) | BDRS |
| Increased Rigidity | (past year) | BDRS |
| Increased egocentricity | (past year) | BDRS |
| Impairment of regard for feelings of others | (past year) | BDRS |
| Coarsening of affect | (past year) | BDRS |
| Impairment of emotional control | For example, increased petulance and irritability (past year) | BDRS |
| Hilarity in inappropriate situations | (past year) | BDRS |
| Diminished emotional responsiveness | (past year) | BDRS |
| Sexual misdemeanor | Arising *de novo* in old age (past year) | BDRS |
| Hobbies relinquished | (past year) | BDRS |
| Diminished Initiative/growing apathy | (past year) | BDRS |
| Purposeless activity | (past year) | BDRS |
| Needs Reminders | Does the patient need reminders or advice to manage chores, do shopping, cooking, play games or handle money? | DS |
| Needs Help to Remember | Does the patient need help to remember important things such as appointments, recent events or names of family members or friends? | DS |
| Needs Help Finding Things | Does the patient need frequent (at least once a month) help finding misplaced objects, keeping appointments or maintaining health or safety (locking doors, taking medication)? | DS |
| Needs Household Chores Done | Does the patient need household chores done for them? | DS |
| Needs Watching When Awake | Does the patient need to be watched or kept company when awake? | DS |
| Needs to be Escorted When Outside | Does the patient need to be escorted when outside? | DS |
| Needs to be Accompanied Bathing/Eating | Does the patient need to be accompanied when bathing or eating? | DS |
| Needs to be Dressed/Washed/Groomed | Does the patient have to be dressed, washed and groomed? | DS |
| Needs to be Taken to Toilet | Does the patient have to be taken to the toilet regularly to avoid incontinence? | DS |
| Needs to be Fed | Does the patient have to be fed? | DS |
| Needs to be Turned/Moved/Transferred | Does the patient need to be turned, moved or transferred? | DS |
| Needs to Wear Diaper/Catheter | Does the patient wear a diaper or a catheter? | DS |
| Needs to be Tube Fed | Does the patient need to be tube fed? | DS |
| Equivalent Institutional Service | Level of care that the patient receives and requires | DS |
| Admission to Hospital | Any admission to a hospital? (past 6 months) | Personal report |
| Treatment | Treatment for any medical condition (past 6 months) | Personal report |
| Had seizure? | Has the patient had a seizure? (past 6 months) | Personal report |
| DELUSIONS | Any delusions? (past month) | CUSPAD |
| HALLUCINATIONS | Any hallucinations? (past month) | CUSPAD |
| ILLUSIONS | Any illusions? (past month) | CUSPAD |
| Agitation | Any agitation or aggression? (past month) | CUSPAD |
| Sadness/Depression | Any sadness or depression? (past month) | CUSPAD |
| Depression Frequency | Depression frequency? (past month) | CUSPAD |
| Appetite | Has the patient's appetite changed? (past month) | CUSPAD |
| Extrapyramidal symptoms | Presence of any extrapyramidal symptoms | UPDRS-III |
| Tremor | Presence of moderate to severe resting tremor | UPDRS-III |
| Bradykinesia | Presence of moderate to severe bradykinesia | UPDRS-III |
| Gait | Presence of moderate to severe gait abnormality | UPDRS-III |
| Myoclonus | Presence of moderate to severe myoclonus | UPDRS-III |
| Rigidity | Presence of moderate to severe rigidity in neck or limbs | UPDRS-III |
| Fluctuating Cognition (Lewy) | Fluctuating cognition? (past year) | FC/DLB |
| Visual Hallucinations (Lewy) | Visual hallucinations? (past year) | FC/DLB |
| Sum of Orientation variables | Sum of orientation section of MMSE | MMSE |
| MMSE -- Repeat three objects | Sum of immediate recall section of MMSE (range = 0-3) | MMSE |
| MMSE -- World | Sum of "Spell the word 'WORLD' backwards" section of MMSE (range = 0-5) | MMSE |
| MMSE -- Memory of three objects | Sum of delayed recall section of MMSE (range = 0-3) | MMSE |
| Sum of Language variables | Sum of language section of MMSE (range = 0-8) | MMSE |
| Intersecting pentagons-MMS | Construction item of MMSE | MMSE |
| Total MMSE Score (range 0-30) | Total MMSE sum score | MMSE |

CUSPAD: The Columbia University Scale for Psychopathology in Alzheimer's disease^1^

MMSE: Mini Mental State Evaluation^2^

BDRS: Blessed Dementia Rating Scale^3^

DS: Dependence Scale^4^

UPDRS-III: Unified Parkinson’s Disease Rating Scale ^5^

FC/DLB: Lewy Body Questionnaire

Input References
